# Slower Learning Rates from Negative Outcomes in Substance Use Disorder over a 1-Year Period and their Potential Predictive Utility

**DOI:** 10.1101/2021.10.18.21265152

**Authors:** Ryan Smith, Samuel Taylor, Jennifer L. Stewart, Salvador M. Guinjoan, Maria Ironside, Namik Kirlic, Hamed Ekhtiari, Evan J. White, Haixia Zheng, Rayus Kuplicki, Tulsa 1000 Investigators, Martin P. Paulus

**Affiliations:** Laureate Institute for Brain Research, Tulsa, OK, USA; Department of Community Medicine, University of Tulsa, Tulsa, OK USA

**Keywords:** Substance Use Disorders, Computational Modeling, Active Inference, Learning Rate, Explore-Exploit Dilemma, Directed Exploration

## Abstract

Computational modelling is a promising approach to parse dysfunctional cognitive processes in substance use disorders (SUDs), but it is unclear how much these processes change during the recovery period. We assessed 1-year follow-up data on a sample of treatment-seeking individuals with one or more SUDs (alcohol, cannabis, sedatives, stimulants, hallucinogens, and/or opioids; *N* = 83) that were previously assessed at baseline within a prior computational modelling study. Relative to healthy controls (HCs; *N* = 48), these participants were found at baseline to show altered learning rates and less precise action selection while completing an explore-exploit decision-making task. Here we replicate these analyses when these individuals returned and re-performed the task 1 year later to assess the stability of these baseline differences. We also examine whether baseline modelling measures can predict symptoms at follow-up. Bayesian analyses indicate that: (a) group differences in learning rates were stable over time (posterior probability = 1); (b) intra-class correlations (ICCs) between model parameters at baseline and follow-up were significant and ranged from small to moderate (.25 ≤ ICCs ≤ .54); and (c) learning rates and/or information-seeking values at baseline were associated with substance use severity at 1-year follow-up in stimulant and opioid users (.36 ≤ *r*s ≤ .43, .002 ≤ *p*s ≤ .02). These findings suggest that learning dysfunctions are moderately stable during recovery and could correspond to trait-like vulnerability factors. In addition, computational measures at baseline had some predictive value for changes in substance use severity over time and could be clinically informative.

## 1. Introduction

Substance use disorders (SUDs) are among the most common, costly, and burdensome psychiatric conditions (NIMH, 2007; Suzuki & Kober, 2018). Despite considerable research to date (Everitt & Robbins, 2016; Valyan, Ekhtiari, Smith, & Paulus, 2020), understanding of the cognitive and neurobiological underpinnings of these conditions remains incomplete, with limited ability to inform treatment or predict symptom change over time. Computational modelling represents a promising approach for further elucidating the neural and cognitive mechanisms underlying SUDs. This approach can account for maladaptive perceptual, learning, and decision-making processes, as well as generate quantitative hypotheses at multiple levels of description. Several computational modelling and neuroimaging studies over the last two decades have found evidence that compulsive behavior patterns seen in SUDs are associated with a shift from so-called ‘model-based’ (goal-directed) to ‘model-free’ (habitual) control (Donamayor, Strelchuk, Baek, Banca, & Voon, 2018; Everitt & Robbins, 2005, 2016; Obst et al., 2018; Reiter et al., 2016; Sebold et al., 2014; Sjoerds et al., 2013; Voon et al., 2015). Other modelling studies have also reported evidence of altered interoception (Smith, Kuplicki, et al., 2020) and altered approach-avoidance processes in SUDs (Smith, Kirlic, Stewart, Touthang, Kuplicki, Khalsa, et al., 2021; Smith, Kirlic, Stewart, Touthang, Kuplicki, McDermott, et al., 2021). These and other types of maladaptive behavior patterns have been linked to relapse as well as several other negative long-term outcomes (Passetti, Clark, Mehta, Joyce, & King, 2008; Verdejo-Garcia, Chong, Stout, Yucel, & London, 2018). As part of the broader field of computational psychiatry (Huys, Maia, & Frank, 2016), the goal of model-based studies has been to identify and measure differences in the information processing mechanisms that underlie such maladaptive patterns, and to examine if they can aid in assessing symptom severity, guiding treatment decisions, predicting treatment outcomes, and evaluating treatment progress, among others (Smith, Taylor, & Bilek, 2021).

This aim of computational psychiatry to inform personalized medicine approaches – via either treatment prediction or assessment of treatment progress – requires that computational measures provide reliable individual difference estimates over time. That is, measures of computational mechanisms should be consistent over time unless true mechanistic changes have occurred. If changes over time instead reflect random influences, their use as assessment tools will be limited (Nair, Rutledge, & Mason, 2020). To address this, the longitudinal stability of computational measures has been the topic of recent studies, with results ranging from poor to excellent estimates of reliability (Brown, Chen, Gillan, & Price, 2020; Chung et al., 2017; Enkavi et al., 2019; Hedge, Bompas, & Sumner, 2020; Moutoussis et al., 2018; Price, Brown, & Siegle, 2019; Shahar et al., 2019; Smith, Kirlic, Stewart, Touthang, Kuplicki, McDermott, et al., 2021). This highly variable pattern of results suggests that there may be significant measurement error and/or that the cognitive processes engaged during many tasks change with repeated performance (e.g., due to learning). Many commonly used computational tasks are also yet to be assessed for longitudinal stability, or for their ability to track or predict changes over time in clinically relevant variables (e.g., symptom levels, physiological states, etc.). There is thus a need for thorough assessment of the longitudinal reliability of a broader range of task measures within computational psychiatry and for further evaluation of their ability to capture information about states vs. traits.

In a recent paper studying SUDs (Smith, Schwartenbeck, et al., 2020), we used a computational modelling approach to analyze behavior on a commonly used three-armed bandit task (Zhang & Yu, 2013), which is designed to measure the balance between information-seeking and reward-seeking during decision-making under uncertainty (i.e., solving the ‘explore-exploit dilemma’; (Addicott, Pearson, Sweitzer, Barack, & Platt, 2017)). This dataset included healthy controls (HCs; *N* = 54) and a community sample of individuals with one or more SUDs (alcohol, cannabis, sedatives, stimulants, hallucinogens, and/or opioids; *N* = 147). This was part of the Tulsa 1000 (T1000) project (Victor et al., 2018) – a naturalistic longitudinal study recruiting subjects based on the dimensional NIMH Research Domain Criteria framework (Insel et al., 2010). Computational modelling in that prior study provided evidence that, relative to HCs, substance users learned more slowly from losses and more quickly from wins. Substance users also showed less precise (less value-sensitive) decisions, corresponding to a behavioral tendency to change decision strategies despite prior success. While these results suggested a mechanism whereby substance users may continue with maladaptive behavior (under uncertainty) despite negative consequences, the stability of these differences was not addressed. Namely, it was not clear whether these results reflected stable trait vulnerability factors, or were dependent on current psychological states, or whether they may track symptom changes over time.

Participants in the T1000 project were invited to return for a 1-year follow-up visit and asked to complete – among other assessments - the above-mentioned three-armed bandit task. This afforded the opportunity to (1) test the individual- and group-level stability of baseline results over time (i.e., whether/how computational phenotypes may change during the recovery process), and (2) examine whether baseline computational measures predict clinical differences at follow-up. This study reports the results of these analyses as a means of examining the clinical utility of this task/model as a potential clinical assessment tool.

## 2. Methods

### 2.1 Participants

Participants represent a subset of those from our original baseline study (Smith, Schwartenbeck, et al., 2020) who agreed to return for a 1-year follow-up visit. In the baseline study, these participants were identified from the exploratory subsample (i.e., first 500 participants) of the T1000 project (Victor et al., 2018), which recruited a community sample of subjects based on the dimensional NIMH Research Domain Criteria framework. The T1000 study included individuals 18-55 years old, screened on the basis of dimensional psychopathology scores: Drug Abuse Screening Test (DAST-10 (Bohn, Babor, & Kranzler, 1991)) score > 3, Patient Health Questionnaire (PHQ-9 (Kroenke, Spitzer, & Williams, 2001)) ≥ 10, and/or Overall Anxiety Severity and Impairment Scale (OASIS (Norman, Hami Cissell, Means-Christensen, & Stein, 2006)) ≥ 8. HCs did not have psychiatric diagnoses or show elevated symptoms. Participants were excluded if they: (a) tested positive for drugs of abuse via urine screen, (b) met criteria for psychotic, bipolar, or obsessive-compulsive disorders, or (c) reported history of moderate-to-severe traumatic brain injury, neurological disorders, severe or unstable medical conditions, active suicidal intent or plan, or change in medication dose within 6 weeks. See (Victor et al., 2018) for a more complete description of inclusion/exclusion criteria.

The study was approved by the Western Institutional Review Board. All participants provided written informed consent prior to completion of the study protocol, in accordance with the Declaration of Helsinki, and were compensated for participation. ClinicalTrials.gov identifier: #NCT02450240.

After baseline screening, participants were grouped based on DSM-IV-TR or DSM-5 diagnosis using the Mini International Neuropsychiatric Inventory (MINI version 6.0 or 7.0) (D. Sheehan et al., 2015; D. V. Sheehan & Lecrubier, 2010; D. V. Sheehan et al., 1998). In our baseline study, we focused on treatment-seeking individuals with SUDs (*N* = 147; including alcohol, cannabis, sedatives, stimulants, hallucinogens, and/or opioid use disorder) with or without comorbid depression and anxiety disorders. These individuals were compared to 54 HCs with no mental health diagnoses. Most substance users were currently enrolled in a residential facility or maintenance outpatient program after completion of more intensive treatments (mean days abstinent = 92; SD = 56). Due to a difference between HCs and SUDs in scores on the Wide Range Achievement Test (WRAT) – a commonly used measure of premorbid IQ (Johnstone, Callahan, Kapila, & Bouman, 1996) – our prior study also confirmed results in a subsample propensity matched on this measure (as well as on age and sex). This included 51 HCs and 49 SUDs. Of the participants who were invited to return for the 1-year follow-up, 48 HCs and 83 substance users agreed to participate (45 HCs and 25 SUDs in the propensity matched subsample). **Table 1** lists group demographics and clinical measures for both the baseline and follow-up samples by group (only including those that returned for follow-up). **Table 2** also lists diagnosis frequency for specific SUDs and anxiety/depression for baseline and follow-up (including all participants, showing that diagnostic composition did not change with dropout).

**Table 1:** Descriptive Statistics (Means and Standard Deviations) for Demographic and Clinical Measures by Group and Session.

| Full Sample |  |  |  |  |  |  |  |  |  |
| --- | --- | --- | --- | --- | --- | --- | --- | --- | --- |
|  | Total | Baseline |  | Follow-up |  | Usable Data (N) | Effect of Clinical Status | Effect of Session | Effect of Clinical Status/Session Interaction |
|  |  | HCs | SUDs | HCs | SUDs |  |  |  |  |
|  | 131 | 48 | 83 | 48 | 83 |  |  |  |  |
| Age | 34.17<br>(10.18) | 32.19<br>(11.19) | 35.32<br>(9.46) | N/A | N/A | HC: 48<br>SUD+: 83<br>Total: 131 | $t(129) = -1.7$<br>$p = 0.09$<br>$d = -0.3$ | N/A | N/A |
| Sex (Male) | 114<br>(44%) | 22<br>(46%) | 35<br>(42%) | N/A | N/A | HC: 48<br>SUD+: 83<br>Total: 131 | $\chi^2(1) = 0.17$<br>$p = 0.68$ | N/A | N/A |

| **DAST** | 3.48<br>(3.72) | 0.12<br>(0.39) | 7.55<br>(2.17) | 0.45<br>(0.55) | 2.72<br>(3.01) | HC: 38<br>SUD+: 81<br>Total: 119 | **F(1, 117) = 193.62**<br>**p < 0.001**<br>**η²=0.62** | **F(1, 117) = 177.72**<br>**p < 0.001**<br>**η²=0.6** | **F(1, 117) = 102.46**<br>**p < 0.001**<br>**η²=0.47** |
| --- | --- | --- | --- | --- | --- | --- | --- | --- | --- |
| **PHQ** | 3.67<br>(5.1) | 0.88<br>(1.33) | 6.84<br>(6.11) | 1.08<br>(1.81) | 3.35<br>(4.69) | HC: 38<br>SUD+: 80<br>Total: 118 | **F(1, 116) = 30.51**<br>**p < 0.001**<br>**η²=0.21** | **F(1, 116) = 25.15**<br>**p < 0.001**<br>**η²=0.18** | **F(1, 116) = 16.44**<br>**p < 0.001**<br>**η²=0.12** |
| **OASIS** | 3.7<br>(4.35) | 1.48<br>(2.01) | 5.99<br>(4.74) | 1.39<br>(2.27) | 3.74<br>(4.49) | HC: 38<br>SUD+: 81<br>Total: 119 | **F(1, 117) = 27.47**<br>**p < 0.001**<br>**η²=0.19** | **F(1, 117) = 177.72**<br>**p < 0.001**<br>**η²=0.6** | **F(1, 117) = 8.82**<br>**p = 0.004**<br>**η²=0.07** |
| **WRAT** | 60.42<br>(6.26) | 63.78<br>(4.61) | 58.45<br>(6.31) | N/A | N/A | HC: 45<br>SUD+: 77<br>Total: 122 | **t(120) = 4.94**<br>**p < 0.001**<br>**d = 0.9** | N/A | N/A |
| **Regular**<br>**Nicotine**<br>**Smoker\*** | 68<br>(26%) | 7<br>(15%) | 27<br>(33%) | N/A | N/A | HC: 46<br>SUD+: 47<br>Total: 93 | **χ²(1) = 17.87**<br>**p < 0.001** | N/A | N/A |
| **Propensity-Matched** |  |  |  |  |  |  |  |  |  |
|  | Total | Baseline |  | Follow-up |  | Usable<br>Data (N) | Effect of Clinical<br>Status | Effect of Session | Effect of Clinical<br>Status/Session<br>Interaction |
|  |  | HCs | SUDs | HCs | SUDs |  |  |  |  |
|  | 70 | 45 | 25 | 45 | 25 |  |  |  |  |
| **Age** | 32.4<br>(10.37) | 32.27<br>(11.25) | 32.65<br>(8.91) | N/A | N/A | HC: 45<br>SUD+: 25<br>Total: 70 | t(68) = -0.14<br>p = 0.89<br>d = -0.04 | N/A | N/A |
| **Sex (Male)** | 68<br>(49%) | 21<br>(47%) | 13<br>(52%) | N/A | N/A | HC: 45<br>SUD+: 25<br>Total: 70 | χ²(1) = 0.18<br>p = 0.67 | N/A | N/A |
| **DAST** | 2.31<br>(3.4) | 0.13<br>(0.4) | 7.6<br>(2.36) | 0.45<br>(0.55) | 3.76<br>(3.37) | HC: 38<br>SUD+: 25<br>Total: 63 | **F(1, 61) = 186.8**<br>**p < 0.001**<br>**η²=0.75** | **F(1, 61) = 24.9**<br>**p < 0.001**<br>**η²=0.29** | **F(1, 61) = 60.05**<br>**p < 0.001**<br>**η²=0.5** |
| **PHQ** | 2.74<br>(4) | 0.87<br>(1.36) | 7.68<br>(5.15) | 1.08<br>(1.81) | 3.72<br>(3.96) | HC: 38<br>SUD+: 25<br>Total: 63 | **F(1, 61) = 63.99**<br>**p < 0.001**<br>**η²=0.51** | **F(1, 61) = 6.59**<br>**p = 0.01**<br>**η²=0.1** | **F(1, 61) = 16.84**<br>**p < 0.001**<br>**η²=0.22** |
| **OASIS** | 3.01<br>(3.8) | 1.44<br>(1.99) | 6.72<br>(4.46) | 1.39<br>(2.27) | 4.56<br>(4.33) | HC: 38<br>SUD+: 25<br>Total: 63 | **F(1, 61) = 40.52**<br>**p < 0.001**<br>**η²=0.4** | F(1, 61) = 2.69<br>p = 0.11<br>η²=0.04 | **F(1, 61) = 5.59**<br>**p = 0.02**<br>**η²=0.08** |
| **WRAT** | 63.29<br>(4.83) | 63.78<br>(4.61) | 62.4<br>(5.22) | N/A | N/A | HC: 45<br>SUD+: 25<br>Total: 70 | t(68) = 1.14<br>p = 0.26<br>d = 0.28 | N/A | N/A |
| Regular<br>Nicotine<br>Smoker* | 32<br>(23%) | 7<br>(16%) | 9<br>(36%) | N/A | N/A | HC: 44<br>SUD+: 17<br>Total: 61 | $\chi^2(1) = 8.69$<br>$p = 0.003$ | N/A | N/A |

\* defined as >3650 lifetime cigarettes. DAST = Drug Abuse Screening Test. PHQ = Patient Health Questionnaire. OASIS = Overall Anxiety Severity and Impairment Scale. WRAT = Wide Range Achievement Test. Significant effects are bolded.

**Table 2:** Lifetime DSM-IV/DSM-5 psychiatric disorders within SUDs.

|  | Full Dataset |  |  | Propensity-Matched |  |  |
| --- | --- | --- | --- | --- | --- | --- |
|  | Baseline<br>(N=147) | Follow-up<br>(N=83) | Analysis | Baseline<br>(N = 49) | Follow-up<br>(N=25) | Analysis |
| **Substance Use Disorders** |  |  |  |  |  |  |
| **Alcohol** | 55 (37%) | 30 (36%) | $\chi^2(1) = 0.04$<br>$p = 0.85$ | 20 (41%) | 10 (40%) | $\chi^2(1) = 0$<br>$p = 0.95$ |
| **Cannabis** | 55 (37%) | 23 (28%) | $\chi^2(1) = 2.23$<br>$p = 0.14$ | 17 (35%) | 5 (20%) | $\chi^2(1) = 1.71$<br>$p = 0.19$ |
| **Stimulants** | 104 (71%) | 61 (73%) | $\chi^2(1) = 0.2$<br>$p = 0.66$ | 35 (71%) | 18 (72%) | $\chi^2(1) = 0$<br>$p = 0.96$ |
| **Opioids** | 56 (38%) | 30 (36%) | $\chi^2(1) = 0.09$<br>$p = 0.77$ | 25 (51%) | 14 (56%) | $\chi^2(1) = 0.16$<br>$p = 0.68$ |
| **Sedatives** | 38 (26%) | 21 (25%) | $\chi^2(1) = 0.01$<br>$p = 0.93$ | 14 (29%) | 9 (36%) | $\chi^2(1) = 0.43$<br>$p = 0.51$ |
| **Hallucinogens** | 5 (3%) | 3 (4%) | $\chi^2(1) = 0.01$<br>$p = 0.93$ | 2 (4%) | 2 (8%) | $\chi^2(1) = 0.5$<br>$p = 0.48$ |
| **2+ Disorders** | 94 (64%) | 51 (61%) | $\chi^2(1) = 0.14$<br>$p = 0.71$ | 34 (69%) | 18 (72%) | $\chi^2(1) = 0.05$<br>$p = 0.82$ |
| **Alcohol Only** | 9 (6%) | 6 (7%) | $\chi^2(1) = 0.11$<br>$p = 0.74$ | 3 (6%) | 2 (8%) | $\chi^2(1) = 0.09$<br>$p = 0.76$ |
| **Cannabis Only** | 9 (6%) | 4 (5%) | $\chi^2(1) = 0.17$<br>$p = 0.68$ | 4 (8%) | 2 (8%) | $\chi^2(1) = 0$<br>$p = 0.98$ |
| **Stimulants Only** | 26 (18%) | 17 (20%) | $\chi^2(1) = 0.27$<br>$p = 0.6$ | 6 (12%) | 3 (12%) | $\chi^2(1) = 0$<br>$p = 0.98$ |
| **Opioids Only** | 8 (5%) | 5 (6%) | $\chi^2(1) = 0.03$<br>$p = 0.85$ | 2 (4%) | 0 (0%) | $\chi^2(1) = 1.05$<br>$p = 0.31$ |
| **Sedatives Only** | 0 (0%) | 0 (0%) | NA | 0 (0%) | 0 (0%) | NA |

| Mood, Anxiety, Stress Disorders |  |  |  |  |  |  |
| --- | --- | --- | --- | --- | --- | --- |
| Major Depressive | 78 (53%) | 43 (52%) | $\chi^2(1) = 0.03$<br>$p = 0.85$ | 30 (61%) | 15 (60%) | $\chi^2(1) = 0.01$<br>$p = 0.92$ |
| Generalized Anxiety | 22 (15%) | 14 (17%) | $\chi^2(1) = 0.15$<br>$p = 0.7$ | 9 (18%) | 7 (28%) | $\chi^2(1) = 0.91$<br>$p = 0.34$ |
| Social Anxiety | 19 (13%) | 11 (13%) | $\chi^2(1) = 0.01$<br>$p = 0.94$ | 8 (16%) | 3 (12%) | $\chi^2(1) = 0.24$<br>$p = 0.62$ |
| Panic | 17 (12%) | 10 (12%) | $\chi^2(1) = 0.01$<br>$p = 0.91$ | 7 (14%) | 3 (12%) | $\chi^2(1) = 0.07$<br>$p = 0.79$ |
| Post-Traumatic Stress | 23 (16%) | 14 (17%) | $\chi^2(1) = 0.06$<br>$p = 0.81$ | 10 (20%) | 5 (20%) | $\chi^2(1) = 0$<br>$p = 0.97$ |
| 2+ Disorders | 46 (31%) | 30 (36%) | $\chi^2(1) = 0.56$<br>$p = 0.45$ | 18 (37%) | 10 (40%) | $\chi^2(1) = 0.08$<br>$p = 0.78$ |

**Note:** Stimulants = amphetamine, methamphetamine, and/or cocaine.

### 2.2 Procedure

T1000 participants underwent a thorough assessment of demographic, clinical and psychiatric factors. The complete list of assessments and supportive references are provided in (Victor et al., 2018). Here we focus on the same symptom measures assessed in the baseline study (i.e., DAST, PHQ, and OASIS).

To address our questions about the longitudinal reliability and predictive utility of computational measures gathered at baseline, participants performed the same three-armed bandit task at follow-up (Zhang & Yu, 2013). This task includes 20 blocks of 16 trials (see top left of **Figure 1**). In each block, participants are told they can choose one of three options on each trial, and that each option has a different probability of reward that doesn’t change throughout the block. They are also informed that the probabilities can change at the start of each new block, but they are not told what the probabilities are. Thus, with each block participants start with no knowledge of the reward probabilities, and they must decide how many times to ‘test out’ each option (‘explore’) before concluding they know which choice has the highest reward probability (i.e., and ‘exploit’ be continuing to choose that option until the end of the block). Reward probabilities were generated from a Beta (2,2) distribution prior to the start of data collection. Identical reward probabilities were used across participants, with pseudorandomized block order.

**Figure 1.**
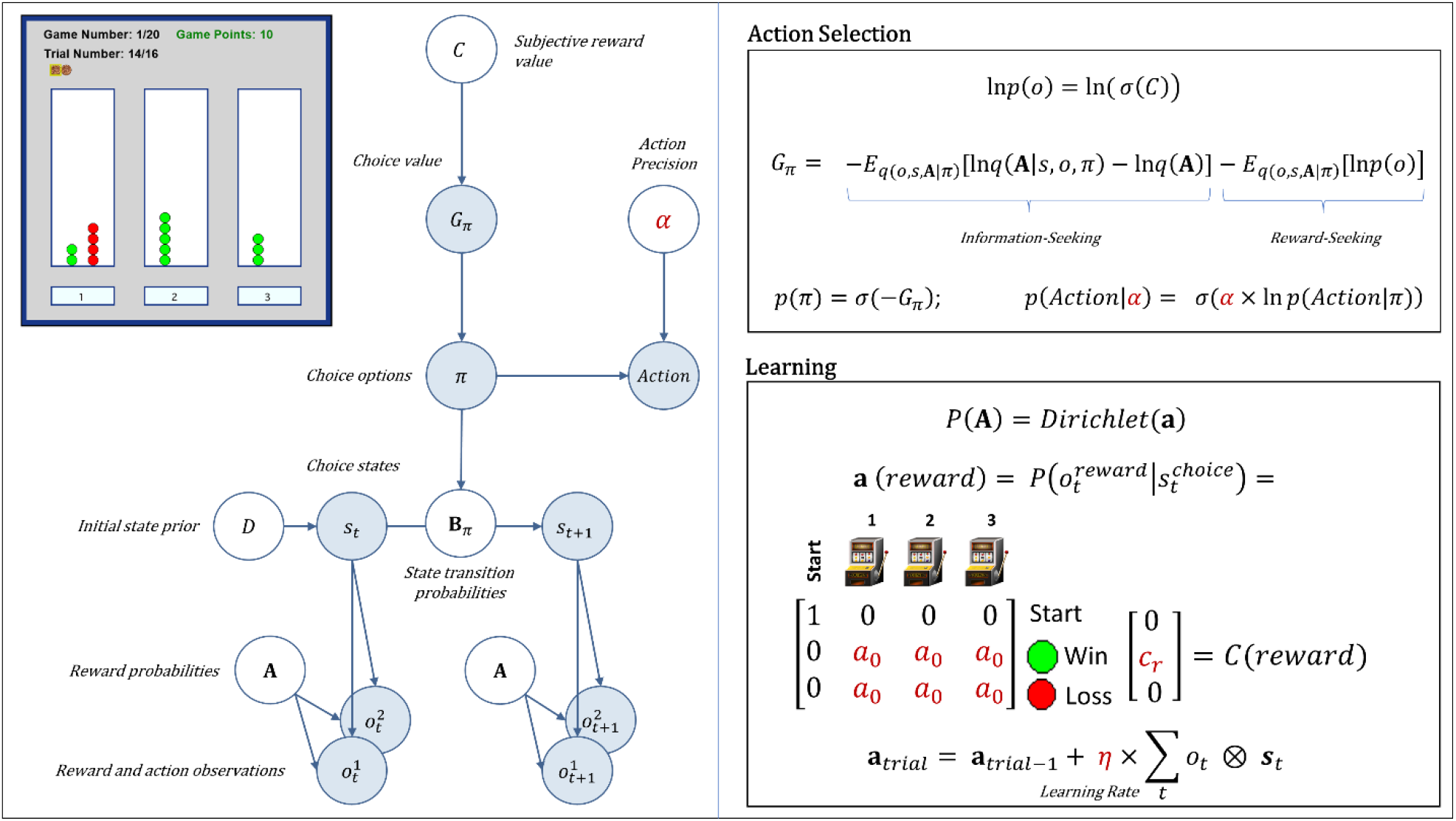
*Upper left*: Illustration of the task interface (for each of three choices, green circle = win; red circle = loss). This task is designed to quantify how individuals switch between an information-seeking and reward-seeking strategy. In each of 20 games, participants had to sample from 3 different choice options with unknown (stable) reward probabilities of winning/losing, with the goal of maximizing reward. The optimal strategy is to start by “exploring” (trying all possible options) to gain information about the probability of winning for each option, and then begin “exploiting” after a few trials by repeatedly choosing the option with highest reward probability. Each game had a known number of trials (16) per game – corresponding to 16 tokens that could be used by pressing one of the 3 buttons (below the white panels on the left, middle and right sides of the interface). After placing each token, they earned 1 point if the token turned green or zero points if the token turned red. Each token decision lasted about 2 sec. After the button press, the chosen option became highlighted for 250ms, after which the token turned green or red to reveal the choice outcome. Participants were instructed to find the most rewarding option and maximize the points earned in each game. Participants were paid an additional $5 or $10 based on task performance. *Left panel*: Graphical depiction of the computational (partially observable Markov decision process) model used with the task. The values of variables in blue circles are inferred on each trial, whereas parameter values in white circles are fixed on each trial. Here, arrows indicate dependencies between variables such that observations 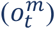 for each modality *m* (reward and observed choice) at a time *t* depend on choice states (*s*_*t*_) at time *t*, where these relationships, 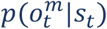, are specified by a matrix **A**. States depend on both previous states and the choice of action policy (*π*), as specified by policy-dependent transition matrices **B**_*π*_ that encode *P*(*s*_*t*+1_*|s*_*t*_, *π*). States at *t* = 1 have an initial state prior specified by a vector *D*. Here, *D* = [1 0 0 0]^*T*^, such that the participant always started in an undecided ‘start’ state at the beginning of each trial. The probability of selecting an action policy depends on its expected free energy (*G*_*π*_), which in turn depends on the subjective reward value of making different observations (e.g., a win vs. loss) for the participant (in a vector *C*). These preferences are defined as a participant’s *log-expectations* over observations, ln*p*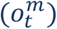. As shown in the top-right panel, the values in *C* are passed through a softmax (normalized exponential) function, *σ*(), which transforms them into a proper probability distribution, and then converted into log probabilities. *Top right panel*: Specifies the mathematical form of the dependencies between *C, G*_*π*_, *π*, and *α* in action selection. When there is no uncertainty about states (as is true of this task), *G*_*π*_ assigns higher values to actions that are expected to simultaneously maximize information gain and reward. The first term on the right corresponds to expected information gain under approximate posterior beliefs (*q*). Large values for this first term indicate the expectation that beliefs about reward probabilities (**A**) will undergo a large change (i.e., that a lot will be learned about these probabilities) given a choice of policy, due to the states and observations it is expected to generate. The second term on the right motivates reward maximization, where a high reward value corresponds to a precise prior belief over a specific observation,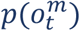. For example, if the subjective value of a win in *C* were *c*_*r*_ = 4 (see bottom left panel), this would indicate a greater subjective reward (higher prior probability) than *c*_*r*_ = 2. The policy expected to maximize the probability of a win (under the associated beliefs about states, observations, and reward probabilities) is therefore favored. Because the two terms in expected free energy are subtracted, policies associated with high expected reward and high expected information gain will be assigned a lower expected free energy. This formulation entails that information-seeking dominates when reward probabilities are uncertain, while reward-seeking dominates when uncertainty is low. A softmax function, *σ*(), then transforms the negative expected free energies into a probability distribution over policies, such that policies with lower expected free energies are assigned higher probabilities. When actions are subsequently sampled from the posterior distribution over policies, randomness in chosen actions is controlled by an action precision parameter (*α*). *Bottom panel*: After each observation of a win/loss, learning corresponds to updating beliefs in a Dirichlet distribution (**a**) over the likelihood matrix **A** that encodes reward probabilities. Here, columns indicate (from left to right) a starting state (pre-choice) and choices 1, 2, and 3, where the rows (from top to bottom) indicate the pre-choice (no reward) observation, observing reward, or no reward. The value of ***a***_0_ – the *insensitivity to information* parameter – is the starting value for beliefs about reward probabilities. These beliefs always start by making up an uninformative (flat) distribution, but higher starting values (e.g., 5 vs. 0.5) effectively down-weight the information-gain term in the expected free energy – leading to an insensitivity to the need for information. The values within **a** (*reward*) are then updated based on the bottom equation, controlled by a learning rate parameter (*η*). For more details regarding the associated mathematics, see the main text and supplemental materials, as well as (Da Costa et al., 2020; K. J. Friston, Lin, et al., 2017; K. J. Friston, Parr, & de Vries, 2017; Smith, Friston, & Whyte, 2021). Estimated model parameters are shown in dark red.

### 2.3 Computational modeling

To model task behavior, we adopted the same partially observable Markov decision process (POMDP) model used at baseline. This approach was motivated by the fact that these models can test for differences in learning rates, random exploration, goal-directed exploration, and sensitivity to information (Schwartenbeck et al., 2019), each of which can contribute to explore/exploit decisions in distinct ways. Estimating the (potentially suboptimal) values of these parameters for each individual can provide insights into the specific decision processes that may promote maladaptive behavior in SUDs (Schwartenbeck et al., 2015). For details about the structure and mathematics of this general class of models, see (Da Costa et al., 2020; Smith, Friston, et al., 2021).

The model is described in full detail in **Supplementary Materials**. Example simulations are also shown in **Supplementary Figure S1**. The model is identical to that used in our previous paper and is outlined in **Table 3**. The model is also depicted graphically (with associated equations) in **Figure 1** and described in detail in the legend. Briefly, the model was defined by (1) the choice states available on each trial in the task, (2) the possible outcomes of those choices (wins/losses), (3) the reward probabilities under each choice state, and (4) the reward value of each possible outcome. Free parameters that influence behavior in the model include: action precision (*α*), reward sensitivity (*c*_*r*_), learning rate (*η*), and insensitivity to information (***a***_0_). The action precision parameter controls the level of stochasticity in choice. Lower values promote choices that are less consistent with beliefs about reward probabilities. In explore-exploit tasks, this corresponds most closely to the construct of random exploration (i.e., choosing actions more randomly as a means of gathering information in the context of high uncertainty). However, random choices in later trials are less consistent with an exploration-based interpretation. The reward sensitivity parameter reflects how much an individual values observing a win. Importantly, as described in **Supplementary Materials**, decision-making is based on a weighted trade-off between expected reward and expected information gain. This means that lower reward sensitivity values will lead individuals to place more value on information-seeking and lead to greater goal-directed exploration. Learning rates quantify how quickly an individual’s beliefs about reward probabilities change when observing each new win/loss. (i.e., influencing how quickly the value of information decreases over time). Insensitivity to information reflects baseline levels of confidence in beliefs about the probability of wins vs. losses for each choice (i.e., before making any observations). Higher insensitivity also leads to reduced goal-directed exploration, because an individual sees less need to seek information a priori. However, unlike reward sensitivity, the influence of this parameter interacts with learning (i.e., higher values also have the effect of making beliefs about reward probabilities less malleable).

**Table 3.**
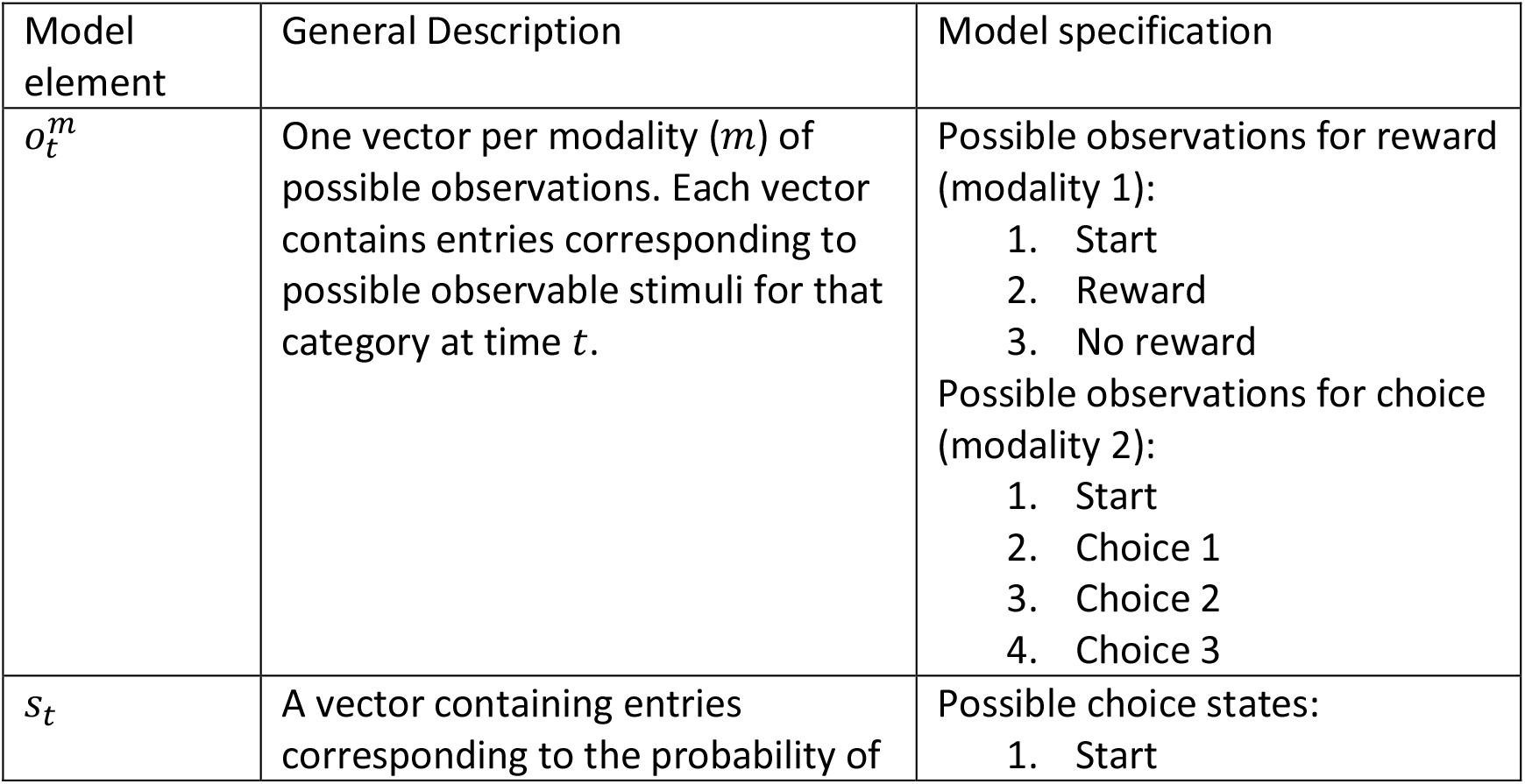
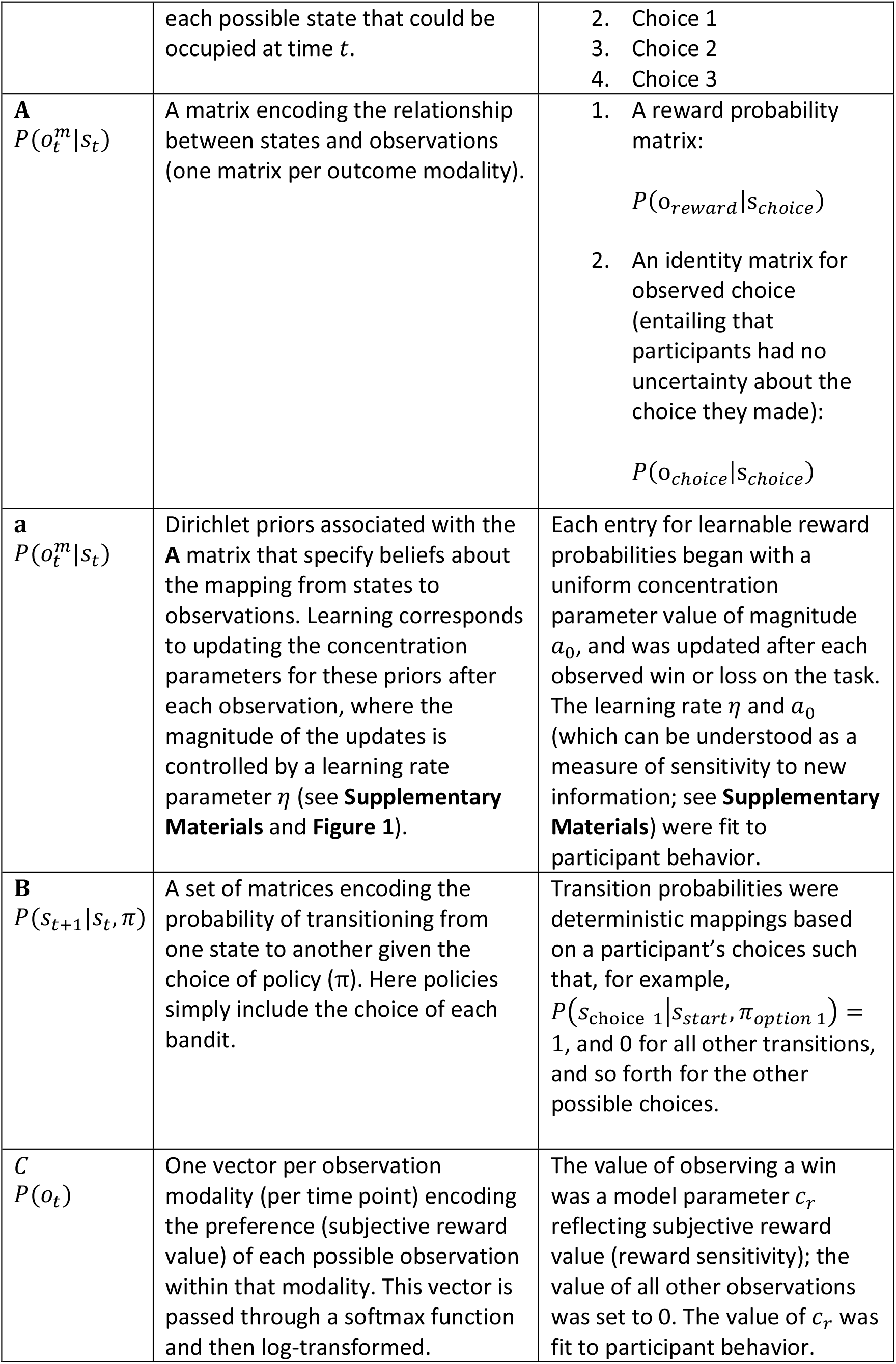
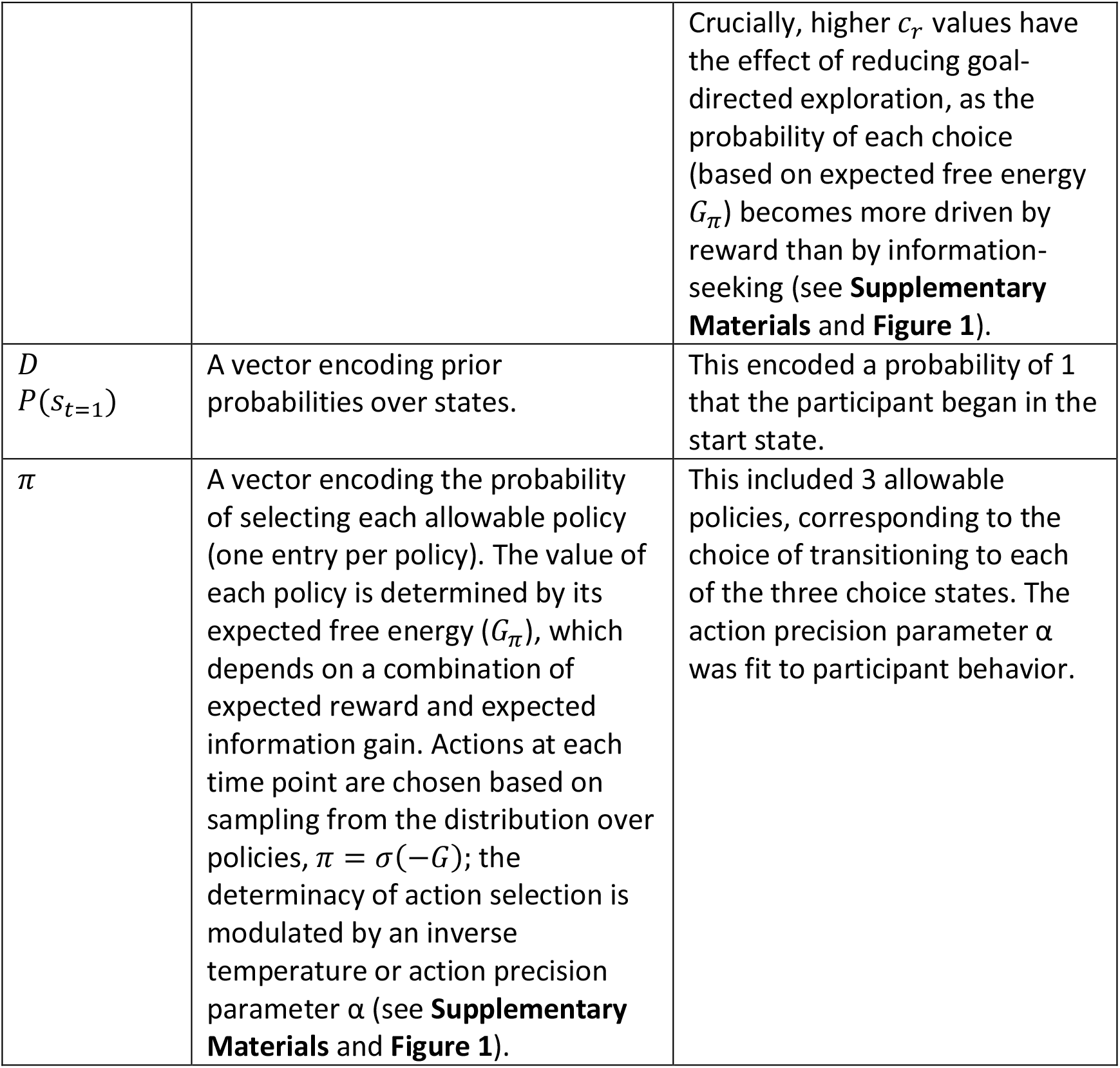
Computational model description.

Estimating these parameters (*α, c*_*r*_, *η*, ***a***_0_) for each individual therefore affords investigation of the mechanisms that can lead to maladaptive choice under uncertainty on an individual basis (Schwartenbeck et al., 2015). Model simulations were run using standard routines available in SPM12 academic software (http://www.fil.ion.ucl.ac.uk/spm/; see software note). As with our prior study, we estimated 10 different nested models, illustrated in **Table 4**, each with different combinations of possible parameters. Bayesian model comparison was then performed to determine the best model (based on (Rigoux, Stephan, Friston, & Daunizeau, 2014; Stephan, Penny, Daunizeau, Moran, & Friston, 2009)). Variational Bayes (variational Laplace; (K. Friston, Mattout, Trujillo-Barreto, Ashburner, & Penny, 2007)) was used to estimate parameter values that maximized the likelihood of each participant’s responses, as described in (Schwartenbeck & Friston, 2016). After establishing the winning model, we confirmed parameter recoverability by simulating behavior under the range of parameter values observed in participants. We then ran the estimation routine on this behavior and examined correlations between the generative and estimated parameters.

**Table 4.** Nested models.

| Parameter: | $\alpha$<br>(action precision) | $c_r$<br>(reward sensitivity) | $\eta$<br>(learning rate) | $a_0$<br>(insensitivity to information) |
| --- | --- | --- | --- | --- |
| Default value if not estimated | 4 | (always estimated) | (removed from model) | 0.25 |
| Prior means during estimation* | 4 | 4 | 0.5 | 0.25 |
| Model 1 | Y | Y | N | N |
| Model 2 | Y | Y | Y | N |
| Model 3 | Y | Y | Y | Y |
| Model 4 | N | Y | Y | Y |
| Model 5 | N | Y | Y | N |
| Model 6 | N | Y | N | N |
| Model 7 | N | Y | N | Y |
| Model 8 | Y | Y | N | Y |
| Model 9** | Y | Y | Wins/Losses | Y |
| Model 10 | Y | Y | Wins/Losses | N |

**Y** indicates that a parameter was estimated for that model; **N** indicates that a parameter was not estimated for that model.
\* Prior variance for all parameters was set to a precise value of $2^{-2}$ in order to deter over-fitting.
\*\*Winning model

### 2.4 Statistical analyses

All analyses were performed in R or MATLAB. We first re-performed the same model assessment measures as in the original paper for the 1-year follow-up data. This included model accuracy metrics, reflecting (1) the average probability of participants’ actions across trials under the model, and (2) the average percentage of trials for which the highest probability action in the model matched the action chosen by participants (i.e., under subject-specific parameter estimates).

We next examined whether participants who did vs. did not return for the follow-up in each group differed in baseline model parameter values, symptom severity, and/or age, sex, or premorbid IQ. As in our prior study, we then ran a parametric empirical Bayes (PEB) analysis (K. J. Friston et al., 2016; Zeidman et al., 2019) using standard MATLAB routines (see software note) to assess stability of group differences over time in both the full and propensity-matched sample. PEB computes group posterior estimates in a general linear model that incorporates posterior variances of individual-level parameter estimates when assessing evidence for group-level models with and without the presence of effects of group and time (and their interaction). A further benefit of this type of hierarchical Bayesian analysis is that it is robust against concerns related to multiple comparisons (Gelman, Hill, & Yajima, 2012; Gelman & Tuerlinckx, 2000). We specifically ran models including age, sex, WRAT (henceforth referred to as premorbid IQ), group (SUDs versus HCs), time, and their interaction as predictor variables. For consistency with frequentist analyses in our baseline study, and with analyses of model-free variables below, supplementary linear mixed effects models (LMEs) with the same predictors were also run for posterior parameter means as point estimates. We also report associated Bayes factors (BFs) for post-hoc comparisons of hypothesized group differences in these models (i.e., JZS Bayes factor analyses with default prior scales in R; BayesFactor package (Morey & Rouder, 2015; Rouder, Morey, Speckman, & Province, 2012)).

In the full sample, we then estimated the longitudinal stability of overall task performance (total wins) and individual parameter estimates between baseline and 1-year follow-up using single-measure consistency intraclass correlations that account for fixed effects across time [ICC(3, 1)]. We chose this ICC measure due to the expectation that time and/or task familiarity could plausibly influence task behavior equivalently across all participants. Although we note that these should not be interpreted as standard test-retest reliability analyses due to the lengthy time period between sessions, where true changes in participant characteristics can plausibly occur, including changes in symptom severity. To address this possibility, we also examined the relationship between pre-post change scores in parameters and pre-post changes in DAST scores, while accounting for age, sex, and premorbid IQ.

Next, in the SUD group, we examined whether parameter values at baseline could predict symptom severity (DAST) scores at 1-year follow-up, before and after accounting for what could be predicted from differences in baseline symptom levels, age, sex, and premorbid IQ. These analyses were performed across all SUDs, as well as when separating individuals with different SUDs (e.g., only including individuals meeting criteria for stimulant use disorder). For these analyses, and the change score analyses above, six participants in the SUDs group were removed due to floor values for DAST at baseline (i.e., due to abstinence prior to study participation), as this prevented the possibility of measurable symptom decreases.

Finally, to confirm relationships seen at baseline between model parameters and model-free metrics of task behavior, we calculated: (a) mean reaction times (RTs; trimmed using an iterative Grubbs test method to remove outliers until a distribution was found which contained no outliers at a threshold of *p* < .01; (Grubbs, 1969)); and (b) number of stays vs. shifts in bandit selection after win and loss outcomes. We examined the relationship between these metrics and our model parameters to gain more insights into the meaning of observed differences. Toward this end, we examined the first and second halves of the games separately (i.e., first 7 choices vs. final 8 choices) to assess periods wherein exploration vs. exploitation would be expected to dominate. To test for consistency with our baseline findings, we also report results of LMEs assessing effects of group and time (and their interaction) on these measures when accounting for age, sex, and premorbid IQ (as well as associated Bayes factors).

As in our prior study, we note here that each of these analyses are considered exploratory, as part of the pre-defined exploratory sample of T1000 participants. Pre-registered analyses will be done to replicate longitudinal results in the confirmatory sample.

## 3. Results

### 3.1 Model comparison and accuracy

When comparing the 10 nested models (**Table 4**), the same model found at baseline – including action precision (*α*), reward sensitivity (*cr*), separate learning rates for wins (*ηwin*) and losses (*η*_*loss*_), and insensitivity to information (***a***0) was the best model (protected exceedance probability = 1). On average, this model accurately predicted true actions on 63% of trials (SD = 11%); SUDs = 62% (SD = 10%), HCs = 64% (SD = 11%). Average probability assigned to true actions by this model was .57 (SD = .11); SUDs = .57 (SD = .11), HCs = .58 (SD = .11). Note that chance accuracy = 1/3. Parameter recoverability analyses showed that generative and estimated parameters for simulated behavior under this model were highly correlated for the range of parameter values observed in our participants: action precision (*r* = .80, *p* < .001), reward sensitivity (*r* = .90, *p* < .001), learning rate for wins (*r* = .91, *p* < .001), learning rate for losses (*r* = .91, *p* < .001), insensitivity to information (*r* = .79, *p* < .001).

### 3.2 Longitudinal stability of group differences

When comparing individuals at baseline in each group who did vs. did not return for follow-up, those who did not return did not significantly differ in age, sex, OASIS, scores, PHQ scores, or DAST scores at baseline (in either the full or matched samples).

**Table 5** presents descriptive statistics for parameters by group. Bayesian (PEB) analyses testing effects on posterior distributions (means and variances) for each parameter also revealed very strong evidence for a number of effects in both the full and matched samples (posterior probability = 1 in all cases). When assessing potential effects of group, time, and their interaction (and accounting for age, sex, and baseline premorbid IQ), the model with the most evidence in both the full and matched samples included a sustained group difference in learning rate for losses from baseline to follow-up (slower in SUDs, see **Figure 2**; for statistical results in analogous LMEs taking a frequentist approach, see **Table 5**). However, these analyses did not support a sustained difference in action precision or learning rate for wins seen in our previous report, or a group difference in any other parameter. There were also effects of time on reward sensitivity (increases over time) and learning rate for losses (decreases over time) in both samples. There were no interactions between group and time for any parameter in the full sample. In contrast, within the matched sample group by time interactions were present in the winning model for reward sensitivity (steeper increase over time in SUDs) and learning rates for wins (decrease over time in SUDs but increase over time in HCs).

There were also effects of age, sex, and premorbid IQ on some parameters. In the full sample: (1) age was negatively associated with action precision and positivity associated with reward sensitivity, (2) learning rate for wins was faster in males, and (3) higher premorbid IQ was associated with slower learning rate for losses. In the matched sample, reward sensitivity was greater in males and premorbid IQ was positively associated with action precision.

**Table 5:** Model Parameters by Group and Session (Means and Standard Deviations) as well as Results of Linear Mixed Effects Model Analyses.

| Full Sample |  |  |  |  |  |  |  |  |  |
| --- | --- | --- | --- | --- | --- | --- | --- | --- | --- |
|  | Total | Baseline |  | Follow-up |  | Usable Data (N) | Effect of Clinical Status | Effect of Session | Effect of Clinical Status/Session Interaction |
|  |  | HCs | SUDs | HCs | SUDs |  |  |  |  |
|  | 131 | 48 | 83 | 48 | 83 |  |  |  |  |
| Action Precision | 2.44 (0.83) | 2.57 (0.92) | 2.2 (0.58) | 2.71 (0.95) | 2.44 (0.86) | HC: 45<br>SUD+: 77<br>Total: 122 | $F(1, 117) = 4.52$<br>$p = 0.04$<br>$\eta^2 = 0.04$ | $F(1, 120) = 4.41$<br>$p = 0.04$<br>$\eta^2 = 0.04$ | $F(1, 120) = 0.67$<br>$p = 0.41$<br>$\eta^2 = 0.01$ |

| **Reward Sensitivity** | 4.45<br>(1.49) | 4.3<br>(1.47) | 4.25<br>(1.5) | 4.5<br>(1.45) | 4.71<br>(1.49) | HC: 45<br>SUD +: 77<br>Total: 122 | $F(1, 117) = 0.98$<br>$p = 0.32$<br>$\eta^2 = 0.01$ | **$F(1, 120) = 12.74$**<br>**$p < 0.001$**<br>**$\eta^2 = 0.1$** | $F(1, 120) = 1.04$<br>$p = 0.31$<br>$\eta^2 = 0.01$ |
| --- | --- | --- | --- | --- | --- | --- | --- | --- | --- |
| **Learning Rate (Wins)** | 0.5<br>(0.13) | 0.47<br>(0.12) | 0.5<br>(0.13) | 0.49<br>(0.13) | 0.51<br>(0.15) | HC: 45<br>SUD +: 77<br>Total: 122 | **$F(1, 117) = 5.41$**<br>**$p = 0.02$**<br>**$\eta^2 = 0.04$** | **$F(1, 120) = 3.86$**<br>**$p = 0.05$**<br>**$\eta^2 = 0.03$** | $F(1, 120) = 0.01$<br>$p = 0.93$<br>$\eta^2 = 0$ |
| **Learning Rate (Losses)** | 0.38<br>(0.15) | 0.43<br>(0.13) | 0.39<br>(0.15) | 0.39<br>(0.15) | 0.35<br>(0.16) | HC: 45<br>SUD +: 77<br>Total: 122 | **$F(1, 117) = 6.45$**<br>**$p = 0.01$**<br>**$\eta^2 = 0.05$** | **$F(1, 120) = 8.68$**<br>**$p = 0.004$**<br>**$\eta^2 = 0.07$** | $F(1, 120) = 0.14$<br>$p = 0.7$<br>$\eta^2 = 0$ |
| **Information Insensitivity** | 0.78<br>(0.28) | 0.72<br>(0.27) | 0.79<br>(0.29) | 0.76<br>(0.31) | 0.82<br>(0.25) | HC: 45<br>SUD +: 77<br>Total: 122 | $F(1, 117) = 3.63$<br>$p = 0.06$<br>$\eta^2 = 0.03$ | $F(1, 120) = 1.54$<br>$p = 0.22$<br>$\eta^2 = 0.01$ | $F(1, 120) = 0.27$<br>$p = 0.6$<br>$\eta^2 = 0$ |
| **Propensity Matched** |  |  |  |  |  |  |  |  |  |
|  | Total | Baseline |  | Follow-up |  | Usable Data (N) | Effect of Clinical Status | Effect of Session | Effect of Clinical Status/Session Interaction |
|  |  | HCS | SUDs | HCS | SUDs |  |  |  |  |
|  | 70 | 45 | 25 | 45 | 25 |  |  |  |  |
| **Action Precision** | 2.51<br>(0.89) | 2.59<br>(0.94) | 2.12<br>(0.58) | 2.67<br>(0.94) | 2.51<br>(0.9) | HC: 45<br>SUD +: 25<br>Total: 70 | $F(1, 65) = 2.43$<br>$p = 0.12$<br>$\eta^2 = 0.04$ | $F(1, 68) = 2.86$<br>$p = 0.1$<br>$\eta^2 = 0.04$ | $F(1, 68) = 1.63$<br>$p = 0.21$<br>$\eta^2 = 0.02$ |
| **Reward Sensitivity** | 4.4<br>(1.51) | 4.23<br>(1.47) | 4.17<br>(1.63) | 4.51<br>(1.49) | 4.72<br>(1.52) | HC: 45<br>SUD +: 25<br>Total: 70 | $F(1, 65) = 0.02$<br>$p = 0.88$<br>$\eta^2 = 0$ | **$F(1, 68) = 4.84$**<br>**$p = 0.03$**<br>**$\eta^2 = 0.07$** | $F(1, 68) = 0.59$<br>$p = 0.45$<br>$\eta^2 = 0.01$ |
| **Learning Rate (Wins)** | 0.49<br>(0.13) | 0.46<br>(0.12) | 0.53<br>(0.1) | 0.49<br>(0.13) | 0.51<br>(0.16) | HC: 45<br>SUD +: 25<br>Total: 70 | $F(1, 65) = 2.56$<br>$p = 0.11$<br>$\eta^2 = 0.04$ | $F(1, 68) = 0.31$<br>$p = 0.58$<br>$\eta^2 = 0$ | $F(1, 68) = 1.48$<br>$p = 0.23$<br>$\eta^2 = 0.02$ |
| **Learning Rate (Losses)** | 0.39<br>(0.15) | 0.43<br>(0.13) | 0.35<br>(0.17) | 0.39<br>(0.14) | 0.34<br>(0.18) | HC: 45<br>SUD +: 25<br>Total: 70 | **$F(1, 65) = 4.32$**<br>**$p = 0.04$**<br>**$\eta^2 = 0.06$** | $F(1, 68) = 1.33$<br>$p = 0.25$<br>$\eta^2 = 0.02$ | $F(1, 68) = 0.65$<br>$p = 0.42$<br>$\eta^2 = 0.01$ |
| **Information Insensitivity** | 0.77<br>(0.29) | 0.73<br>(0.27) | 0.82<br>(0.33) | 0.75<br>(0.28) | 0.86<br>(0.28) | HC: 45<br>SUD +: 25<br>Total: 70 | $F(1, 65) = 3.23$<br>$p = 0.08$<br>$\eta^2 = 0.05$ | $F(1, 68) = 0.35$<br>$p = 0.56$<br>$\eta^2 = 0.01$ | $F(1, 68) = 0.07$<br>$p = 0.8$<br>$\eta^2 = 0$ |

\*Analyses are reported using results from LMEs accounting for age, sex, and premorbid IQ (WRAT). Significant effects are bolded.

**Figure 2.**
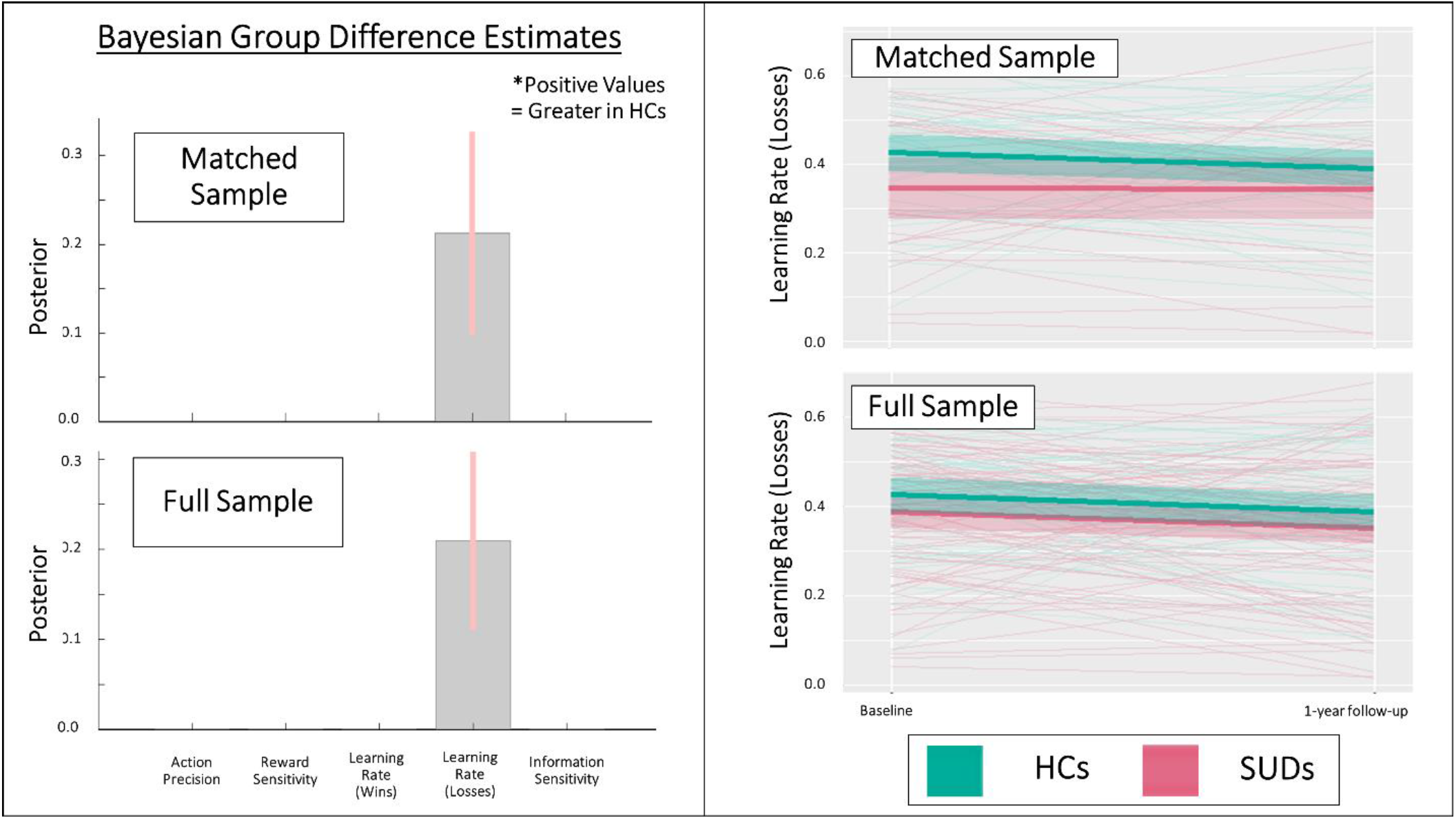
*Left*: Results of parametric empirical Bayes (PEB) analyses, showing the posterior means and variances for group difference estimates in the full and propensity-matched samples in models accounting for age, sex, and premorbid IQ. These Bayesian group comparisons confirm the differences in learning rates for losses seen at baseline. There was also a main effect of time on this learning rate, but no significant interactions between group and time, indicating the group effects were stable. No other parameters showed main effects of group. See main text for further results of these analyses. Learning rate values are in logit-space. *Right*: Spaghetti plots showing individual changes from baseline to follow-up, as well as group means and standard errors, for learning rate for losses in the full and matched samples. HCs = healthy controls, SUDs = substance use disorders.

Additional PEB analyses focused only on 1-year follow-up data (i.e., analogous to those reported in our baseline study, accounting for age, sex, and premorbid IQ) also showed positive evidence for the group difference in learning rate for losses in both the full sample (posterior probability = .83) and matched sample (posterior probability = .93). For plots of each parameter by group and time in both samples, see **Supplementary Figure S2**. For plots of the additional PEB results (illustrating effect sizes) not shown in **Figure 2**, see **Supplementary Figure S3**. For consistency with frequentist analyses in our baseline study, **Table 5** also presents effects of group, session, and their interaction within LMEs predicting the posterior parameter means (with the same additional predictors as the PEB models). Findings were largely consistent with the Bayesian results. However, significant group effects were also present in action precision and learning rate for wins in the full sample (mirroring our previously reported baseline results). Linear models equivalent to those in our baseline paper also supported PEB results in showing significantly slower learning rates for losses in SUDs than HCs when only comparing groups at follow-up (full sample: *t*(117) = 2.137, *p* = .03, *d* =0.40), but showed no other significant differences for other parameters.

### 3.3 Individual-level parameter stability

The ICCs for task performance and parameters between baseline and 1-year follow-up were poor to moderate (see **Table 6** and **Figure 3**), with the highest values across all participants for reward sensitivity (ICC = .54) and learning rate for losses (ICC = .43).With the exception of action precision and total wins, SUDs tended to have numerically higher ICCs than HCs. Task performance (total wins) showed the lowest stability over time across participants (ICC = .15), driven by a non-significant association between baseline and follow-up in the SUD group.

**Table 6.**
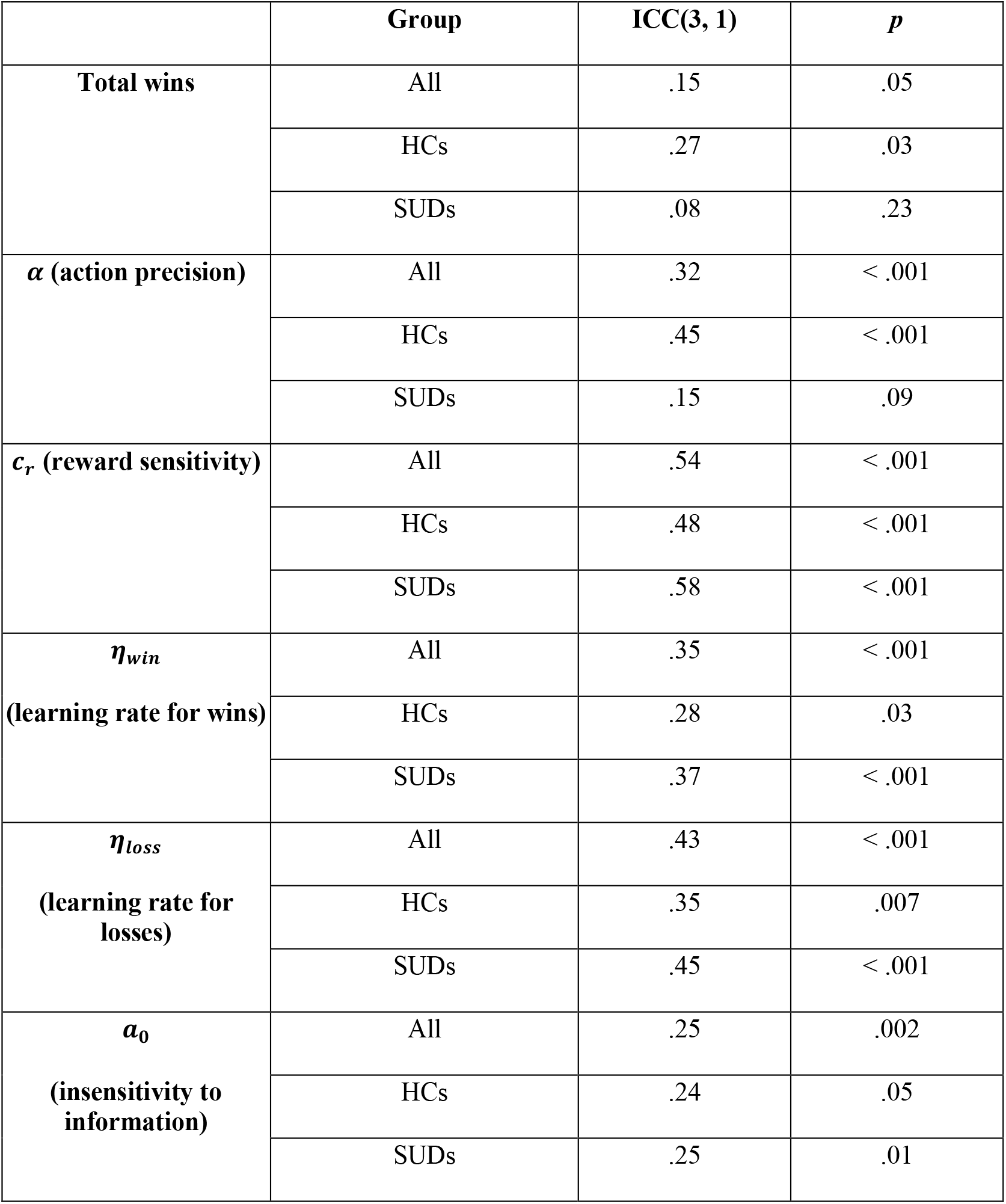
Intra-class correlations between baseline and 1-year follow-up (full sample)

**Figure 3.**
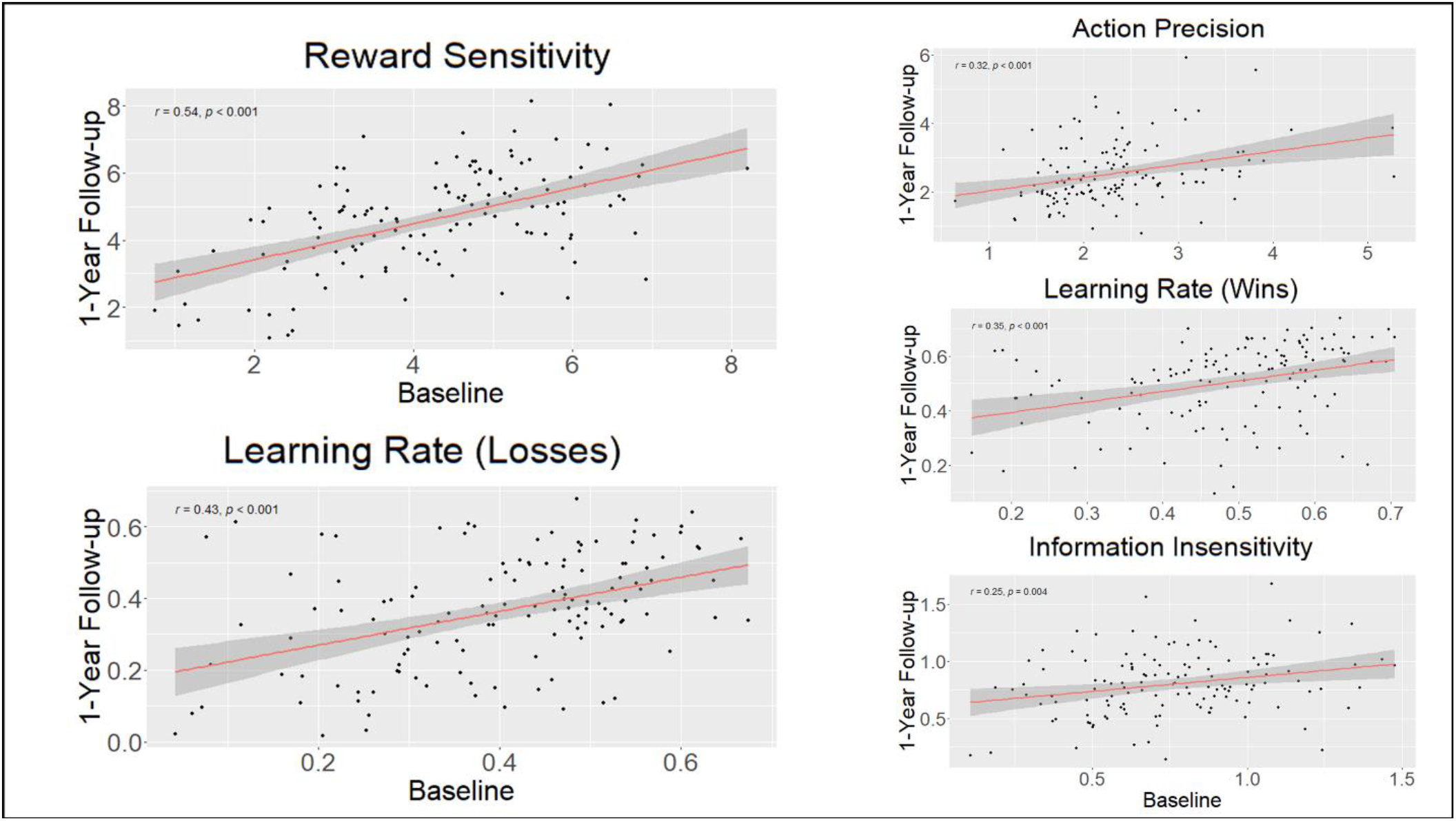
Correlations between computational parameters at baseline and 1-year follow-up.

There were no significant associations between pre-post changes in DAST scores and pre-post changes in parameters across all SUDs. When examining stimulant users and opioid users separately, in both cases there was an association between pre-post changes in DAST scores and pre-post changes in action precision. In stimulant users this correlation was *r* = -.28 (*p* = .03), and this remained unchanged after accounting for the relationship between DAST changes and age, sex, and premorbid IQ scores (*r* = -.29, *p* = .03; see **Figure 4**). In opioid users this correlation was *r* = -.34 (*p* = .07), and this became significant after accounting for the relationship between DAST changes and age, sex, and premorbid IQ scores (*r* = -.38, *p* = .046). No other associations were found.

**Figure 4.**
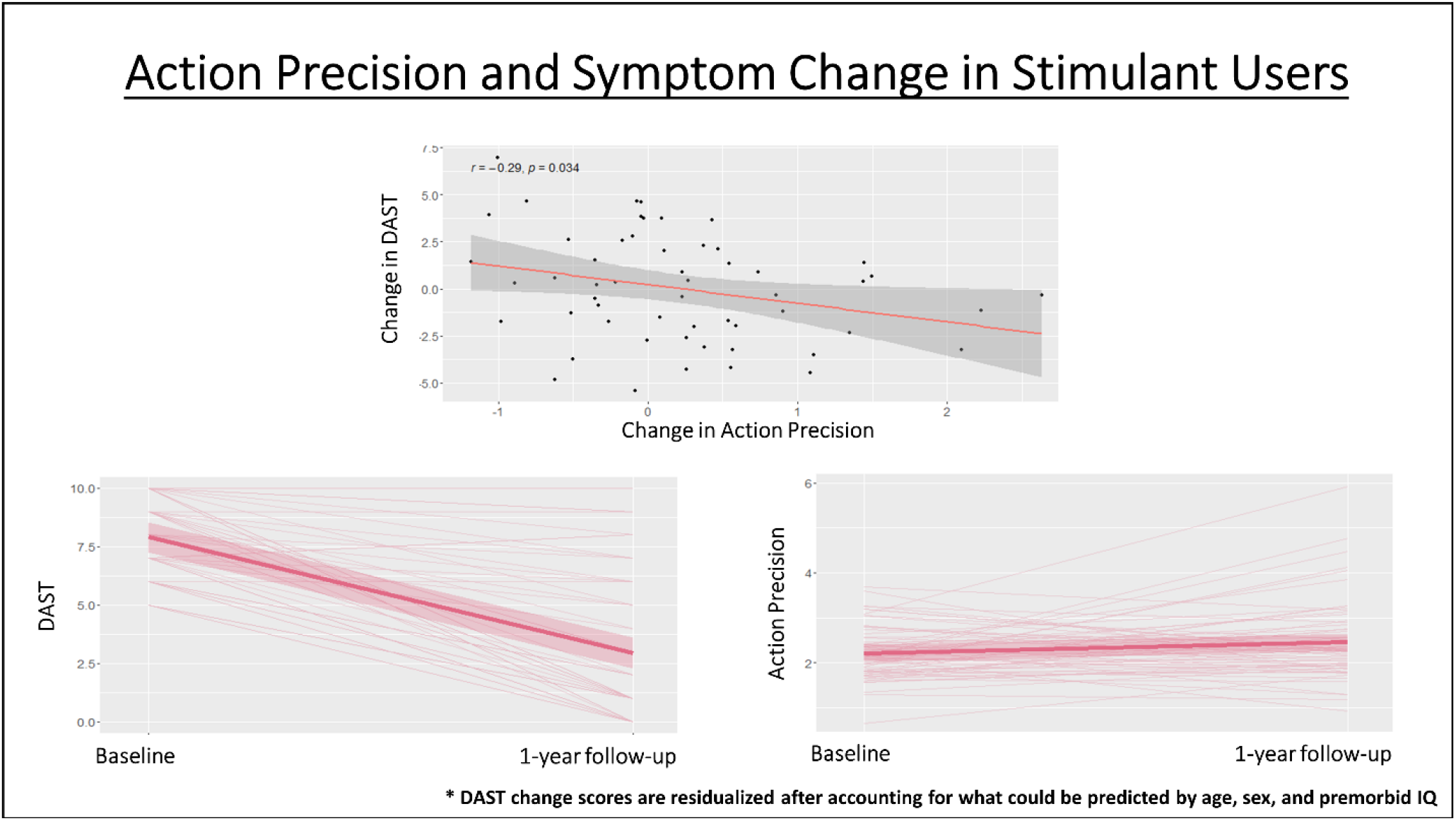
*Top*: Negative correlation in stimulant users (full sample) between pre-to-post changes in action precision and pre-to-post changes in symptom severity (DAST). *Bottom*: Illustration of individual pre-to-post changes in DAST scores and action precision (as well as group mean and SE). As can be seen, DAST scores tend to decrease and action precision tends to increase, but with notable individual differences in each. DAST change scores account for what could already be predicted based on age, sex, and premorbid IQ.

### 3.4 Symptom Change Prediction

In the full sample of substance users, no significant predictive relationships were found between baseline model parameters and DAST scores at 1-year follow-up (after accounting for baseline DAST scores, with or without accounting for age, sex, and premorbid IQ). When restricting analyses to stimulant users, we observed a significantly positive predictive relationship between baseline learning rates for losses and DAST scores at 1-year follow-up (*r* = .33, *p* = .01), which became stronger after accounting for what could be predicted by age, sex, and premorbid IQ (*r* = .4, *p* = .002; see **Figure 5**). Significant negative predictive relationships were also found with baseline learning rates for wins (*r* = -.29, *p* = .03) and insensitivity to information (*r* = -.36, *p* = .005), which each also became stronger after accounting for age, sex, and premorbid IQ (respectively: *r* = -.36, *p* = .007; *r* = -.38, *p* = .004; see **Figure 5**). When restricting analyses to opioid users, we observed a significantly negative predictive relationship between baseline information insensitivity and DAST scores at 1-year follow-up (*r* = -.43, *p* = .02), which weakened after accounting for what could be predicted by age, sex, and premorbid IQ (*r* = -.35, *p* = .07). When restricting analyses to alcohol users, there was a trending negative relationship with information insensitivity (*r* = -.37, *p* = .07), which weakened after accounting for what could be predicted by age, sex, and premorbid IQ (*r* = -.26, *p* = .22). Other specific SUDs were not examined because sample size was considered too low.

**Figure 5.**
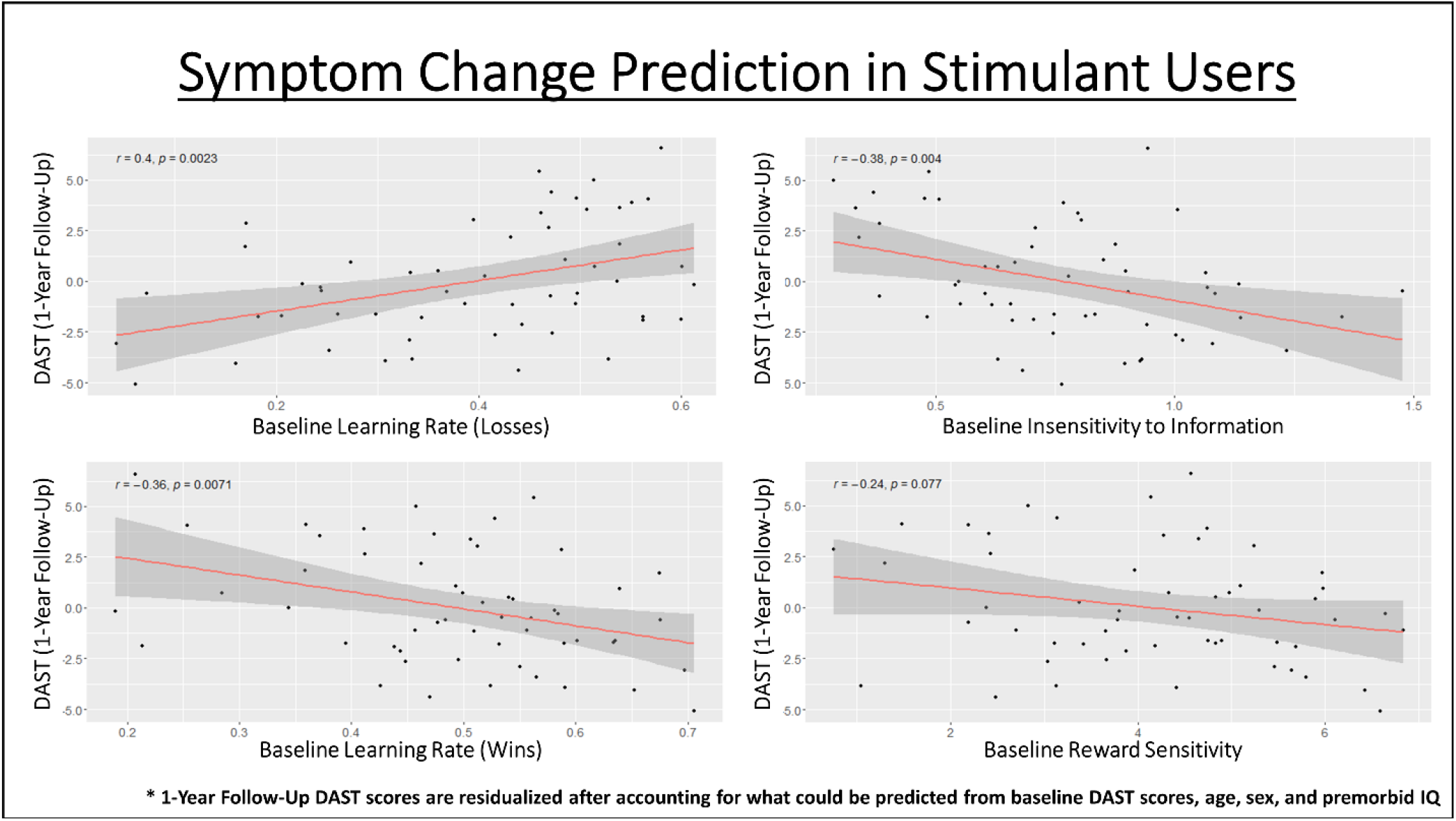
Predictive relationships in stimulant users (full sample) between baseline model parameters and symptom severity at 1-year follow-up, after accounting for what could already be predicted based on age, sex, and premorbid IQ.

### 3.5 Comparison to model-free measures

**Table 7** lists descriptive statistics by group and time in model-free behavioral measures (total wins, win/lose stay/shift choices, and RTs). This table also shows results of LMEs assessing the main effects and interactions between group and time, while accounting for age, sex, and premorbid IQ. In **Supplementary Tables S1-2**, results are further divided into sets derived from early trials (i.e., where information-seeking should be high; choices 2-7 per game), and late trials (i.e., where reward-seeking would be expected to dominate; subsequent 8 choices). Most notably, these results together indicated that, relative to HCs, SUDs showed a larger number of lose-stay choices across time (driven by choices in early trials) in both the propensity-matched and full samples. They also showed a smaller number of lose-shift choices across time (present in both early and late trials) in the full sample.

**Table 7:**
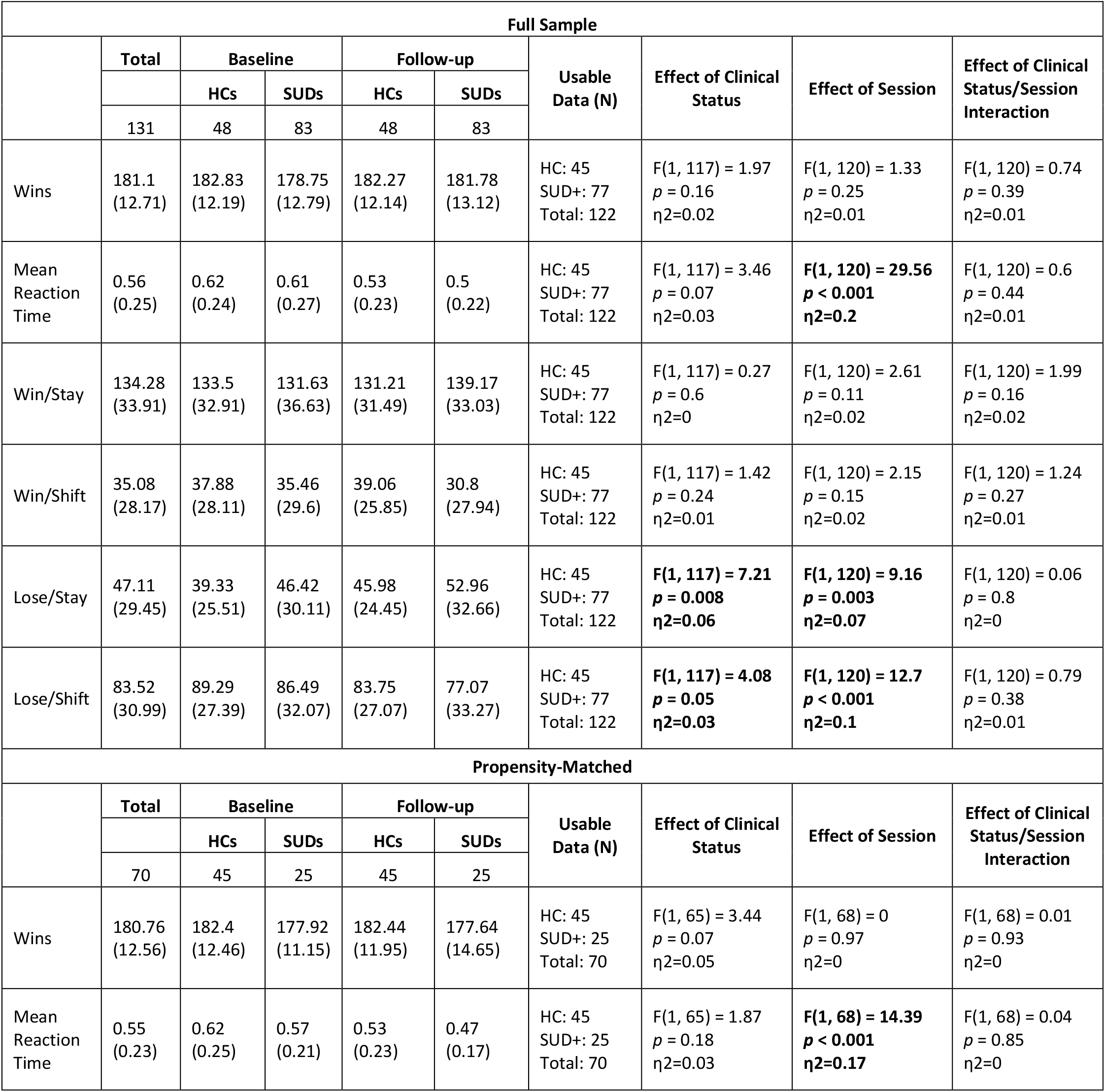
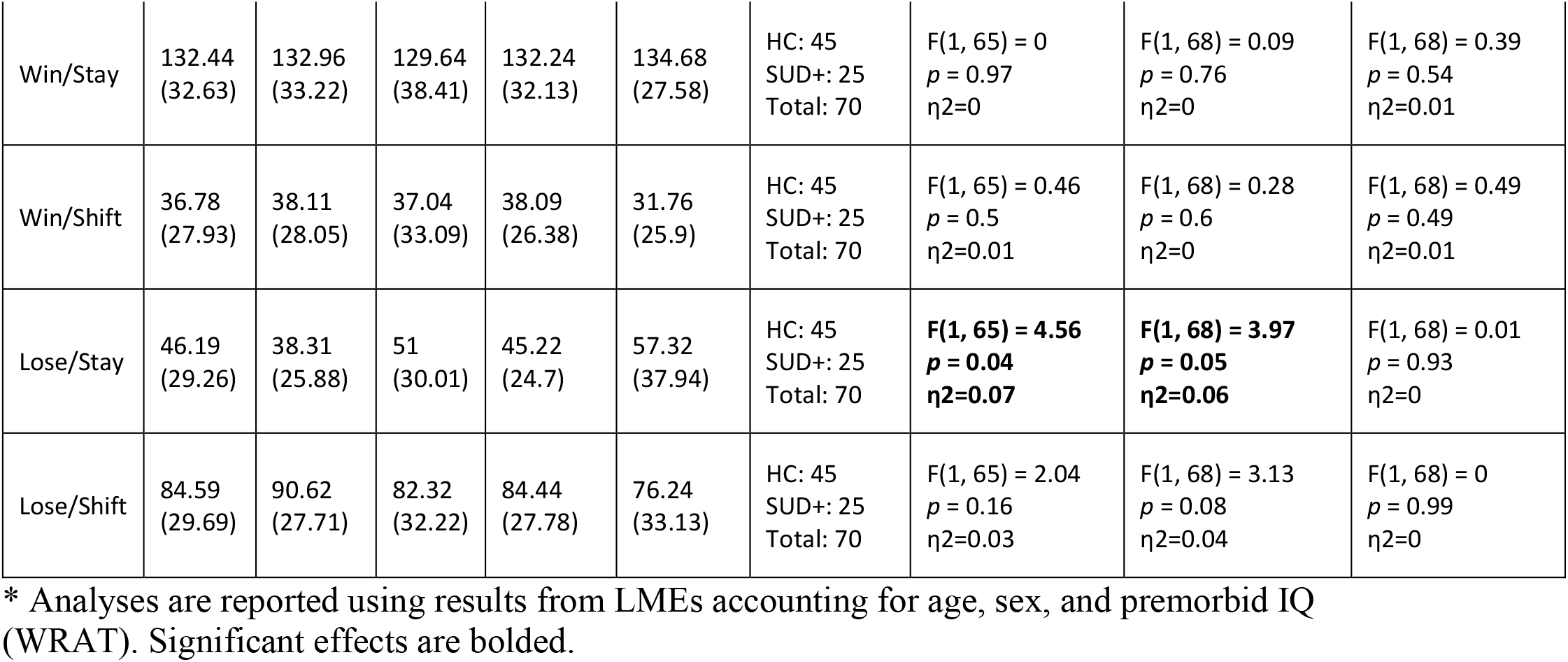
Model Free Task Measures by Group and Session (Means and Standard Deviations)

**Supplementary Figure S4** shows the correlations between model parameters and model-free measures at 1-year follow-up. As can be seen there, results strongly resembled those previously found in our baseline study, First, there was a complex pattern of relationships with win/lose stay/shift behavior in which reward sensitivity and information insensitivity promoted stay behaviors generally, action precision promoted win-stay choices on late trials, and learning rates had relationships with all types of choices in expected directions, but with the strongest relationship to stays vs. switches on loss trials. Number of wins only showed associations with reward sensitivity and action precision (positive relationship). This relationship was notably (numerically) stronger on late trials in each game. RTs were faster in those with higher reward sensitivity, information sensitivity, and learning rate for wins, and slower in those with higher learning rate for losses (*p*s < .001 and BFs > 100 in all cases).

## 4. Discussion

In this study we evaluated the longitudinal stability of both individual- and group-level differences between HCs and SUDs in computational measures of learning and decision-making over a 1-year period. We also examined whether these computational measures could predict changes in symptom severity over time. At the group level, both Bayesian and frequentist analyses showed that a slower learning rate for losses in SUDs (previously observed at baseline (Smith, Schwartenbeck, et al., 2020)) was stable over the 1-year period. Comparison to descriptive measures suggested that this (in part) tracked the fact that SUDs tended to continue with the same decision strategy after incurring a loss (primarily on early trials). This appears consistent with previous results showing associations between SUDs and difficulty avoiding punishment (Myers et al., 2017), diminished responses to negative stimuli (Hester, Bell, Foxe, & Garavan, 2013; Simons & Arens, 2007; Simons, Dvorak, & Batien, 2008; Stewart et al., 2014), reduced sensitivity to losses (Ahn et al., 2014), and a lower impact of large losses on future choices (Petry, Bickel, & Arnett, 1998). Importantly, it could help explain why substance use continues despite negative life consequences. As changes in this learning rate did not correspond to symptom changes over time, it might more plausibly act as a pre-existing (trait) vulnerability factor. For example, those with a greater tendency to persist in a pattern of behavior despite negative outcomes could be more likely to engage in substance use a sufficient number of times to promote addiction. On the other hand, substance misuse over time could lead to less sensitivity to negative outcomes regardless of future symptom change.

At the individual level, we found that some parameters showed moderate stability while others showed poor stability. The two most (moderately) stable parameters were learning rate for losses and reward sensitivity. As the former reflected the primary group differentiator, this further supports its potential role as a pre-existing vulnerability factor, which could act as an adjunct assessment of risk independent of self-report. While parameter estimation error could partly account for these attenuated relationships, we also examined whether the lower levels of stability we observed might be due to associations with individual differences in symptom changes. While not present across all SUDs, in stimulant and opioid users we found that larger reductions in symptom severity were associated with larger increases in action precision, which could suggest this parameter reflects evolving aspects of the disease process. In our baseline study, SUDs showed significantly lower action precision than HCs, while this difference was no longer present at follow-up. This was due to increased action precision over time in SUDs – mirroring the overall reduction in symptom severity at follow-up. Given this pattern, future research should assess whether action precision might act as an objective measure of treatment progress.

When evaluating the predictive utility of baseline parameters, we did not find significant results across all SUDs. However, we did observe significant predictive relationships when restricting analyses to specific SUDs. Namely, we found that symptom severity at follow-up in stimulant users was predicted by baseline learning rate for losses (positive relationship), and also by information insensitivity and learning rate for wins (negative relationships). Opioid users’ symptoms at follow-up showed a similar negative relationship with baseline information insensitivity. If replicated in an independent sample, assessment of these measures at treatment onset might therefore offer additional information about which patients will be more resistant to improvement over time. This represents another important topic for future research.

Despite SUDs showing slower learning from losses (and some evidence for faster learning from wins) at the group level, stimulant users with the slowest learning rates from losses (and fastest learning rates from wins) had better outcomes at follow-up. Also, despite (numerically) greater insensitivity to information in SUDs at the group level, both stimulant and opioid users with the greatest insensitivity also had lower symptoms at follow-up. One might speculate that, upon initiating abstinence, a slower learning rate from negative consequences could attenuate avoidance (akin to reducing lose/switch decisions) of the uncomfortable aspects of the recovery process (e.g., withdrawal, reflection on poor like circumstances in therapy, etc.) and allow a person to persist through a difficult situation without resorting to maladaptive coping mechanisms. However, such possibilities would require further investigation. Greater information insensitivity is also theoretically associated with reduced subjective uncertainty and greater confidence in expected action outcomes. In the right (e.g., therapeutic) circumstances, this could perhaps also play a role in facilitating recovery. However, there are also plausible ways in which these differences might be expected to have opposing effects as well. Independent of their predictive value, future research should therefore further address the theoretical significance and correct interpretation of these relationships, as they could speak to important components of decision-making mechanisms in SUDs that deserve attention as possible targets of behavioral interventions (Verdejo-Garcia et al., 2018; Verdejo-Garcia, Garcia-Fernandez, & Dom, 2019).

Although representative of the population (and therefore potentially more informative in real-world clinical settings), one limitation of this study is the heterogeneity of our SUD group. Several secondary analyses in our baseline study addressed some related concerns, but they nonetheless constrain interpretability here. For example, the predictive relationships we found separately in stimulant and opioid users suggest that other SUDs (e.g., cannabis, sedatives) may have had confounding effects; but samples of individuals with each of these disorders in isolation would be needed to definitively answer this question. Another issue is that, although we did not identify differences in those who did versus did not return for the follow-up visit, drop-out nonetheless reduced the statistical power available for our analyses and could still limit the generalizability of our results. We plan to address these issues further in the confirmatory dataset presently set aside to replicate these results.

In summary, we found that individuals with SUDs showed stable reductions in learning from losses relative to HCs over a 1-year period. Individual-level parameter stability was poor-to-moderate, and in some cases appeared to be attenuated by symptom changes. Finally, multiple model parameters at baseline showed potential predictive utility with respect to symptom changes over time. These results hold promise in the development of adjunct computational assessment tools for predicting symptom evolution and perhaps treatment progress, which could inform treatment decisions.

## Data Availability

All data produced in the present study are available upon reasonable request to the authors

## Software Note

All model simulations, model comparison, and parametric empirical Bayes analyses were implemented using standard routines (**spm_MDP_VB_X.m, spm_BMS.m, spm_dcm_peb.m, spm_dcm_peb_bmc.m**) that are available as MATLAB code in the latest version of SPM academic software: http://www.fil.ion.ucl.ac.uk/spm/. For the specific code used to build the three-armed bandit task model and fit parameters to data, see: https://github.com/rssmith33/3-armed_bandit_task_model.

## Funding

This work has been supported in part by The William K. Warren Foundation, the National Institute of Mental Health (R01MH123691 (RS and RLA); R01MH127225 (RS and SSK); K99MD015736 (EJS)), the National Institute on Drug Abuse (R01DA050677 (JLS)), and the National Institute of General Medical Sciences (P20GM121312 (RS and MPP)).

## Conflict of Interest

None of the authors have any conflicts of interest to disclose.

## Authors’ contribution

RS selected and performed all analyses and wrote the original draft of the manuscript. ST assisted with analyses and edited the manuscript. JLS assisted with analyses and edited the manuscript. SMG, MI, NK, HE, EJW, and HZ edited the manuscript. RK contributed to data processing and edited the manuscript. MPP designed and oversaw the Tulsa 1000 study and edited the manuscript. The Tulsa 1000 investigators each contributed to overseeing the Tulsa 1000 study and edited the manuscript.

## Supplementary Materials

### Computational modeling details

To model behavior on the task, we adopted a Markov decision process (MDP) model under the active inference framework (see main text **Figure 1**); for more details about the structure and mathematics of this class of models, see (Friston et al., 2017a; Friston et al., 2017b; Parr and Friston, 2017; Smith et al., 2021). This approach requires creating a model with specific sets of possible observations 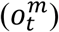, hidden states (*s*_*t*_) that cause those observations, and available actions (policies; *π*). In our model, there were two types of observations (modalities; *m*) that could be made at each time point (*t*). In the first modality 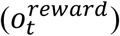, the participant could make a “starting” observation, and then observe either a win or a loss. In the second modality 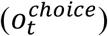, the participant could observe the action that was chosen. Hidden states in the model included a “starting” state as well as the state of having chosen each of the three options). Policies included the three available choices on each trial.

The dependencies between these variables are described by sets of matrices. One set of matrices **A** encodes the way hidden states generate observations, 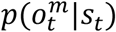. In our model, **A** defines the probability of observing a win vs. a loss given the state of having chosen each option:

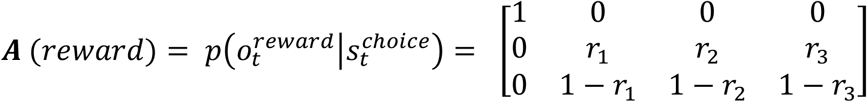

Here, columns indicate (from left to right) the starting state and choices 1, 2, and 3; the rows (from top to bottom) indicate the starting observation, observing a win, or observing a loss. The values of *r*_1_, *r*_2_, *r*_3_ are the true reward probabilities for each choice. There was also a second **A**-matrix mapping each choice state to the observation of that choice, which was set as an identity matrix (i.e., there was no uncertainty in the choice a participant made) and the observation that each choice has been made.

A set of matrices **B**_*π*_ encode state transition probabilities under each policy, *p*(*s*_*t*+1_*|s*_*t*_, *π*). In our model, these defined the transition from the “starting state” to the state of having chosen each possible option under each respective policy. Here, the transition probabilities were simply a deterministic mapping based on participants’ choices, such that, for example, *p*(*s*_*choice* 1_|*s*_*start*_, *π*_*choice* 1_) = 1 and 0 for all other transitions.

A set of vectors *C* encode the subjective reward value of each observation in each modality at each time point. In our model, a value of 0 was fixed for all observations except for observing a win. The value for observing a win was estimated based on participant behavior as an index of reward sensitivity (*c*_*r*_):

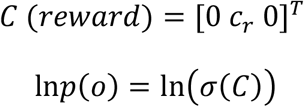

The subjective reward values for different observations are formally specified in terms of a participant’s *log-expectations*. The symbol *σ* indicates a softmax (normalized exponential) function that first transforms the values in *C* into a proper probability distribution, such that higher values for *c*_*r*_ are formally assigned higher prior probabilities (corresponding to greater subjective rewardingness of a win). This distribution is then converted into log probabilities. Higher values of *c*_*r*_ reduce information-seeking (by effectively increasing the weight of the reward-seeking term in expected free energy; shown below).

A vector *D* = [1 0 0 0]^*T*^specified a prior over initial states, such that the participant always started in an undecided starting state at the beginning of each trial.

Action policies (*π*) are assigned value based on a quantity called expected free energy (*G*). When there is no uncertainty about choice states (i.e., no uncertainty about one’s choice on a trial), as is true in our task, the expected free energy can be written as:

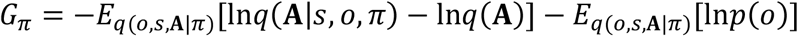

This quantity assigns higher values to actions that are expected to simultaneously maximize information gain and reward. The first term on the right corresponds to information gain. Note that the variable *q*() is used to denote the participant’s (approximate posterior) beliefs in this term. Large values for this first term indicate the expectation that beliefs about reward probabilities will undergo a large change (i.e., that a lot will be learned about these probabilities) given a choice of policy. The second term on the right motivates reward-seeking, by maximizing ln*p*(*o*). Because these terms are subtracted, policies associated with high expected reward and high expected information gain will be assigned a lower expected free energy. This can also be seen more explicitly when expected free energy is shown in the following equivalent form that is cast in terms of model variables (or a full derivation, see (Da Costa et al., 2020)):

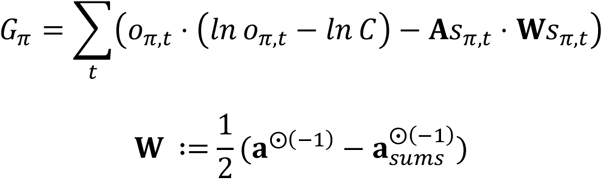

In the first equation it can be seen that policies will have higher value if 1) they minimize the divergence between predicted and preferred outcomes – *o*_*π,t*_. (*ln o*_*π,t*_ - *ln C*) – which can be thought of as maximizing reward probability; and if 2) they seek out states expected to provide the most informative observations about the reward probabilities **– A**s_*π,t*_. **W**s_π,t_ **–** which can be thought of as goal-directed information-seeking. In the second equation, the variable **a** within **W** denotes the current concentration parameters of Dirichlet priors over reward probabilities associated with the **A** matrix. The := symbol indicates that two things are defined to be equivalent, and the ⨀ symbol indicates the element-wise power (i.e., separately raising each element in a matrix to the power of some number). The term **a**_*sums*_ is a matrix of the same size as **a** where each entry within a column corresponds to the sum of the values of the associated column in **a**. At the start of each game, **a** is as follows:

:

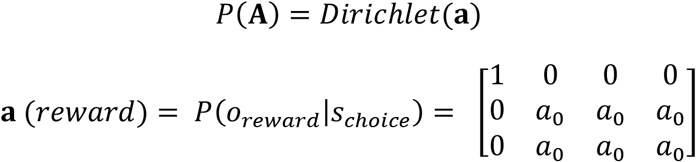

The value of *a*_0_ – the *insensitivity to information* parameter – is the starting value for beliefs about these reward probabilities. These beliefs always start by making up an uninformative (flat) distribution, but higher starting values (e.g., 5 vs. 0.5) effectively down-weight the information-gain term in the expected free energy – leading to an insensitivity to the need for information. Put another way, when information insensitivity (*a*_0_) is high, the need for information-seeking is low a priori. This parameter was estimated for each participant.

The values within **a** (*reward*) are then updated as follows:

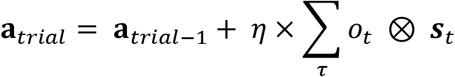

Here ⊗ indicates the cross-product and ***s*** is the posterior belief over choice states (i.e., the belief about which option was chosen). The variable *η* is the *learning rate*, which controls the magnitude of updates in **a** after each observation. This rate can also differ for different observations. Here there were separate learning rates for observing wins vs. losses. A higher learning rate will tend to promote a faster switch to reward-seeking behavior.

Once expected free energy is evaluated, the probability of selecting a policy is:

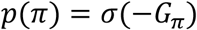

Here, a softmax function (*σ*) transforms the negative expected free energies into a proper probability distribution, such that policies with lower expected free energies are assigned higher probabilities.

A final parameter pertains to choice stochasticity. Active inference naturally distinguishes between uncertainty reduction due to goal-directed, strategic information-seeking (driven by the information gain term in expected free energy) and that due to stochastic choice. The latter approach to gaining information through stochastic choice can be accounted for with an *action precision* parameter (*α*):

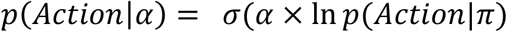

Lower values of *α* increase the probability of selecting actions that disagree with beliefs about the optimal policy.

Based on our model, there are therefore several free parameters that could influence participant behavior: action precision (*α*), reward sensitivity (*c*_*r*_), learning rate (*η*), and the starting value for concentration parameters at the beginning of each game (***a***_0_; henceforth referred to as insensitivity to information). Lower values of *α* produce more randomness in behavior, which could be associated with random exploration (primarily if on early trials). Lower values for *c*_*r*_ and ***a***_0_ produce greater directed exploration in different ways. Higher learning rates promote faster switches from exploration to exploitation. To arbitrate between different model choices, we estimated 10 different nested models – each with different choices in what model variables were included (or fixed at default values) and which to estimate. **Table 4** in the main text shows each model, as well as the default values used for each parameter if not estimated. Note that, based on our primary interest in goal-directed exploration vs. exploitation, *c*_*r*_ was always estimated. We then performed Bayesian model comparison (based on (Rigoux et al., 2014; Stephan et al., 2009)) to determine the best model.

For a tutorial introduction to this general modelling approach, see (Smith et al., 2021); for its implementation within our model, see the **spm_MDP_VB_X.m** MATLAB routine, freely available within the DEM (dynamic expectation maximization) toolbox of the most recent versions of SPM academic software (http://www.fil.ion.ucl.ac.uk/spm/). To illustrate the effect of the information value term in *G*_*π*_, in **Supplementary Figure S1** we show example simulations comparing full model performance to a model where the information value term has been removed. Example simulations under different parameter settings are also shown in **Supplementary Figure S1**.

## Supplementary Tables and Figures

**Table S1:** Model Free Task Measures in Full Dataset by Group and Session (Means and Standard Deviations) split by early (choices 2-7 per block) and late (choices 8-16) trials.

|  | First/Second Half | Total | Baseline |  | Follow-up |  | Usable Data (N) | Effect of Clinical Status | Effect of Session | Effect of Clinical Status/Session Interaction |
| --- | --- | --- | --- | --- | --- | --- | --- | --- | --- | --- |
|  |  |  | HCS | SUDs | HCS | SUDs |  |  |  |  |
|  |  | 131 | 48 | 83 | 48 | 83 |  |  |  |  |
| Wins | First Half | 87.79<br>(7.39) | 89.21<br>(6.75) | 87.13<br>(7.61) | 88.31<br>(6.97) | 87.34<br>(7.75) | HC: 45<br>SUD+: 77<br>Total: 122 | F(1, 117) = 2.83<br>p = 0.1<br>$\eta^2=0.02$ | F(1, 120) = 0.24<br>p = 0.62<br>$\eta^2=0$ | F(1, 120) = 0.08<br>p = 0.78<br>$\eta^2=0$ |
| | Second Half | 93.31<br>(8.11) | 93.62<br>(8.12) | 91.61<br>(8.25) | 93.96<br>(7.41) | 94.45<br>(8.21) | HC: 45<br>SUD+: 77<br>Total: 122 | F(1, 117) = 0.53<br>p = 0.47<br>$\eta^2=0$ | **F(1, 120) = 5.62**<br>**p = 0.02**<br>**$\eta^2=0.04$** | F(1, 120) = 1.21<br>p = 0.27<br>$\eta^2=0.01$ |
| Win/Stay | First Half | 58.61<br>(17.46) | 57.98<br>(18.2) | 58.34<br>(17.67) | 55.48<br>(17.76) | 61.05<br>(16.59) | HC: 45<br>SUD+: 77<br>Total: 122 | F(1, 117) = 0.32<br>p = 0.57<br>$\eta^2=0$ | F(1, 120) = 0.79<br>p = 0.38<br>$\eta^2=0.01$ | F(1, 120) = 1.54<br>p = 0.22<br>$\eta^2=0.01$ |
| | Second Half | 75.68<br>(18.75) | 75.52<br>(17.41) | 73.29<br>(20.55) | 75.73<br>(18.15) | 78.12<br>(17.93) | HC: 45<br>SUD+: 77<br>Total: 122 | F(1, 117) = 0.19<br>p = 0.67<br>$\eta^2=0$ | **F(1, 120) = 4.02**<br>**p = 0.05**<br>**$\eta^2=0.03$** | F(1, 120) = 1.7<br>p = 0.2<br>$\eta^2=0.01$ |
| Win/Shift | First Half | 17.56<br>(15.25) | 19.46<br>(16.07) | 17.25<br>(15.31) | 21.17<br>(15.17) | 14.7<br>(14.41) | HC: 45<br>SUD+: 77<br>Total: 122 | F(1, 117) = 1.98<br>p = 0.16<br>$\eta^2=0.02$ | F(1, 120) = 1.77<br>p = 0.19<br>$\eta^2=0.01$ | F(1, 120) = 1.96<br>p = 0.16<br>$\eta^2=0.02$ |
| | Second Half | 17.52<br>(14.73) | 18.42<br>(14.28) | 18.2<br>(15.53) | 17.9<br>(14.62) | 16.1<br>(14.38) | HC: 45<br>SUD+: 77<br>Total: 122 | F(1, 117) = 0.74<br>p = 0.39<br>$\eta^2=0.01$ | F(1, 120) = 1.78<br>p = 0.18<br>$\eta^2=0.01$ | F(1, 120) = 0.35<br>p = 0.55<br>$\eta^2=0$ |
| Lose/Stay | First Half | 20.69<br>(14.54) | 16.54<br>(11.91) | 21.05<br>(14.9) | 18.12<br>(12.39) | 24.2<br>(15.97) | HC: 45<br>SUD+: 77<br>Total: 122 | **F(1, 117) = 9.83**<br>**p = 0.002**<br>**$\eta^2=0.08$** | **F(1, 120) = 5.57**<br>**p = 0.02**<br>**$\eta^2=0.04$** | F(1, 120) = 0.52<br>p = 0.47<br>$\eta^2=0$ |
| | Second Half | 26.43<br>(16.18) | 22.79<br>(14.98) | 25.37<br>(16.12) | 27.85<br>(14.43) | 28.76<br>(17.59) | HC: 45<br>SUD+: 77<br>Total: 122 | **F(1, 117) = 4.43**<br>**p = 0.04**<br>**$\eta^2=0.04$** | **F(1, 120) = 11.26**<br>**p = 0.001**<br>**$\eta^2=0.09$** | F(1, 120) = 0.05<br>p = 0.82<br>$\eta^2=0$ |
| Lose/Shift | First Half | 43.14<br>(14.56) | 46.02<br>(12.41) | 43.36<br>(14.71) | 45.23<br>(13.17) | 40.05<br>(15.91) | HC: 45<br>SUD+: 77<br>Total: 122 | **F(1, 117) = 5.66**<br>**p = 0.02**<br>**$\eta^2=0.05$** | **F(1, 120) = 3.96**<br>**p = 0.05**<br>**$\eta^2=0.03$** | F(1, 120) = 0.6<br>p = 0.44<br>$\eta^2=0$ |
| | Second Half | 40.38<br>(18.33) | 43.27<br>(17.35) | 43.13<br>(18.86) | 38.52<br>(16.98) | 37.02<br>(18.71) | HC: 45<br>SUD+: 77<br>Total: 122 | F(1, 117) = 2.53<br>p = 0.11<br>$\eta^2=0.02$ | **F(1, 120) = 18.6**<br>**p < 0.001**<br>**$\eta^2=0.13$** | F(1, 120) = 0.66<br>p = 0.42<br>$\eta^2=0.01$ |

\* Significant effects are bolded.

**Table S2:** Model Free Task Measures in Matched Dataset by Group and Session (Means and Standard Deviations) split by early (choices 2-7 per block) and late (choices 8-16) trials.

|  | First/Second Half | Total | Baseline |  | Follow-up |  | Usable Data (N) | Effect of Clinical Status | Effect of Session | Effect of Clinical Status/Session Interaction |
| --- | --- | --- | --- | --- | --- | --- | --- | --- | --- | --- |
|  |  |  | HCS | SUDs | HCS | SUDs |  |  |  |  |
|  |  | 70 | 45 | 25 | 45 | 25 |  |  |  |  |
| Wins | First Half | 88.07<br>(7.29) | 89.31<br>(6.96) | 88.24<br>(6.02) | 88.49<br>(6.81) | 84.92<br>(9.15) | HC: 45<br>SUD+: 25<br>Total: 70 | F(1, 65) = 3.6<br>p = 0.06<br>$\eta^2=0.05$ | F(1, 68) = 2.02<br>p = 0.16<br>$\eta^2=0.03$ | F(1, 68) = 0.98<br>p = 0.32<br>$\eta^2=0.01$ |
| | Second Half | 92.69<br>(7.76) | 93.09<br>(8.09) | 89.68<br>(6.82) | 93.96<br>(7.48) | 92.72<br>(8.21) | HC: 45<br>SUD +: 25<br>Total: 70 | F(1, 65) = 1.89<br>p = 0.17<br>$\eta^2=0.03$ | F(1, 68) = 2.28<br>p = 0.14<br>$\eta^2=0.03$ | F(1, 68) = 0.92<br>p = 0.34<br>$\eta^2=0.01$ |
| Win/Stay | First Half | 58.13<br>(17.32) | 58.2<br>(18.06) | 58.56<br>(18.44) | 56.96<br>(17.35) | 59.68<br>(15.54) | HC: 45<br>SUD +: 25<br>Total: 70 | F(1, 65) = 0.14<br>p = 0.71<br>$\eta^2=0$ | F(1, 68) = 0.02<br>p = 0.88<br>$\eta^2=0$ | F(1, 68) = 0.2<br>p = 0.66<br>$\eta^2=0$ |
| | Second Half | 74.31<br>(17.82) | 74.76<br>(17.61) | 71.08<br>(21.16) | 75.29<br>(18.37) | 75<br>(13.84) | HC: 45<br>SUD +: 25<br>Total: 70 | F(1, 65) = 0.16<br>p = 0.69<br>$\eta^2=0$ | F(1, 68) = 0.56<br>p = 0.46<br>$\eta^2=0.01$ | F(1, 68) = 0.48<br>p = 0.49<br>$\eta^2=0.01$ |
| Win/Shift | First Half | 18.24<br>(15.1) | 19.29<br>(15.73) | 17.76<br>(16.57) | 20<br>(14.79) | 13.64<br>(12.69) | HC: 45<br>SUD +: 25<br>Total: 70 | F(1, 65) = 1.56<br>p = 0.22<br>$\eta^2=0.02$ | F(1, 68) = 0.23<br>p = 0.64<br>$\eta^2=0$ | F(1, 68) = 1.17<br>p = 0.28<br>$\eta^2=0.02$ |
| | Second Half | 18.54<br>(15.02) | 18.82<br>(14.49) | 19.28<br>(17.47) | 18.09<br>(15.05) | 18.12<br>(14.15) | HC: 45<br>SUD +: 25<br>Total: 70 | F(1, 65) = 0<br>p = 0.95<br>$\eta^2=0$ | F(1, 68) = 0.23<br>p = 0.64<br>$\eta^2=0$ | F(1, 68) = 0.01<br>p = 0.91<br>$\eta^2=0$ |
| Lose/Stay | First Half | 19.91<br>(14.54) | 16.29<br>(12.13) | 23.2<br>(15.34) | 18.18<br>(12.4) | 26.28<br>(18.8) | HC: 45<br>SUD +: 25<br>Total: 70 | **F(1, 65) = 6.77**<br>**p = 0.01**<br>**$\eta^2=0.09$** | F(1, 68) = 1.82<br>p = 0.18<br>$\eta^2=0.03$ | F(1, 68) = 0.11<br>p = 0.74<br>$\eta^2=0$ |
| | Second Half | 26.28<br>(16.11) | 22.02<br>(15.1) | 27.8<br>(15.37) | 27.04<br>(14.43) | 31.04<br>(20.13) | HC: 45<br>SUD +: 25<br>Total: 70 | F(1, 65) = 2.44<br>p = 0.12<br>$\eta^2=0.04$ | **F(1, 68) = 5.69**<br>**p = 0.02**<br>**$\eta^2=0.08$** | F(1, 68) = 0.22<br>p = 0.64<br>$\eta^2=0$ |
| Lose/Shift | First Half | 43.72<br>(14.15) | 46.22<br>(12.71) | 40.48<br>(14.91) | 44.87<br>(13.41) | 40.4<br>(16.57) | HC: 45<br>SUD +: 25<br>Total: 70 | F(1, 65) = 3.53<br>p = 0.06<br>$\eta^2=0.05$ | F(1, 68) = 0.22<br>p = 0.64<br>$\eta^2=0$ | F(1, 68) = 0.1<br>p = 0.75<br>$\eta^2=0$ |
| | Second Half | 40.86<br>(17.63) | 44.4<br>(17.3) | 41.84<br>(18.09) | 39.58<br>(16.93) | 35.84<br>(18.58) | HC: 45<br>SUD +: 25<br>Total: 70 | F(1, 65) = 0.95<br>p = 0.33<br>$\eta^2=0.01$ | **F(1, 68) = 7**<br>**p = 0.01**<br>**$\eta^2=0.09$** | F(1, 68) = 0.08<br>p = 0.78<br>$\eta^2=0$ |

\* Significant effects are bolded.

**Figure S1.**
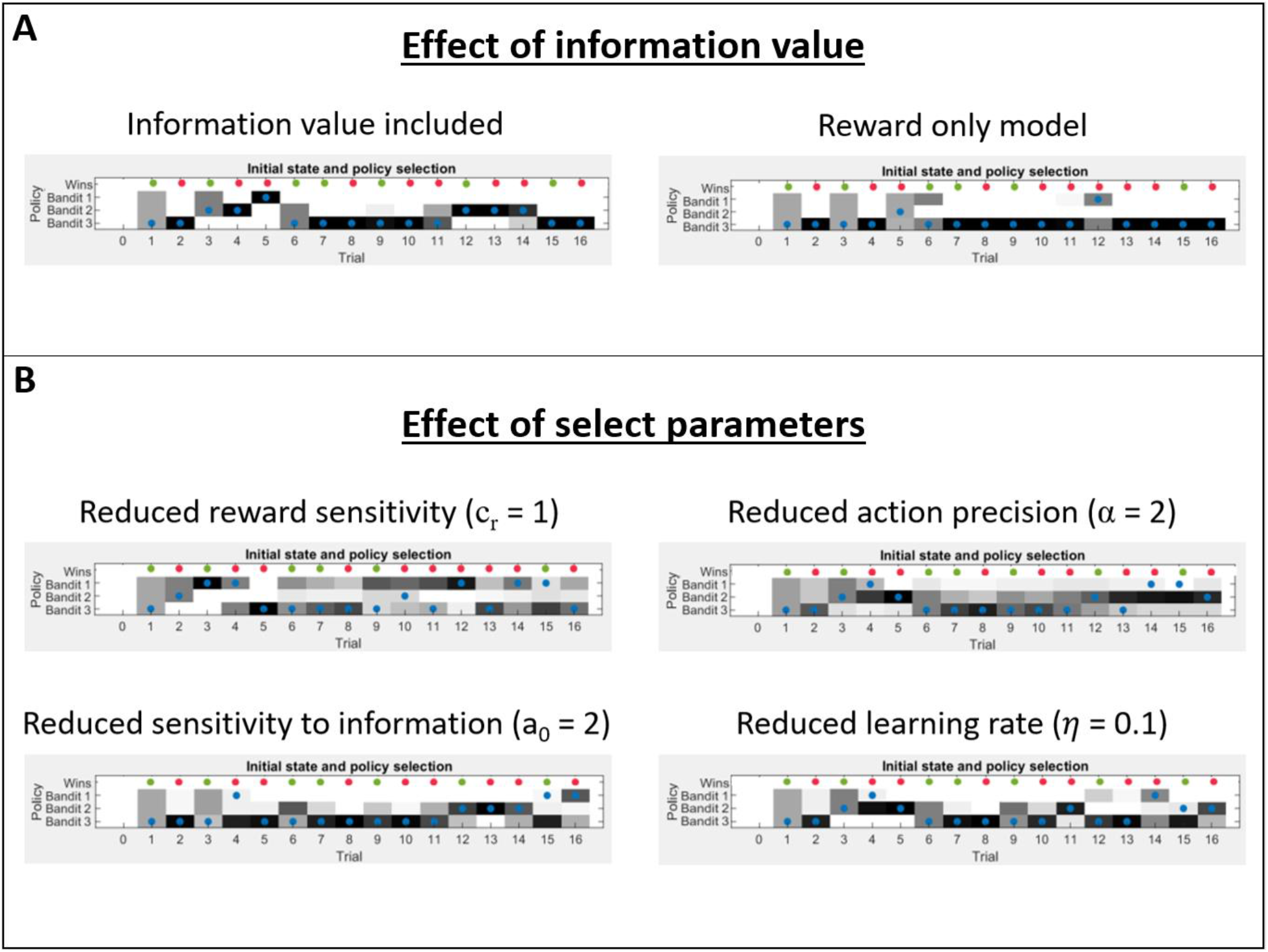
**(A)** Example model simulation of one game with and without the information value term included in policy valuation. Reward probabilities for bandits 1-3 in this game are 0.46, 0.49, and .64 (respectively). Darker shades indicate higher choice probabilities; blue circles indicate the action taken; red and green circles indicate losses and wins (respectively). While the agent on the left panel is driven by both reward maximization and uncertainty reduction, the agent on the right panel only cares about reward. This induces subtle differences in predictions for behavior that are visible, for example, at time step three. Here, after having observed one rewarding and one non-rewarding outcome in bandit three, the agent on the left now prefers to minimize uncertainty about the other two bandits, whereas the agent on the right equally prefers the three bandits because they all have a reward value of 0.5. Here, action precision (AP; lower values promoting random exploration) was set to a high value of 16 to highlight the effects of goal-directed exploration. Reward sensitivity (RS; lower values promote goal-directed exploration) was set to 4, learning rates were set to 0.5, and the prior concentration parameters in the observation model, governing sensitivity to information, were defined as 0.25. **(B)** Example model simulations under single changes from the above-stated parameter values. As can be seen here, reduced RS leads to over-exploration, while reduced sensitivity to information leads to behavior similar to the reward only model. Reduced AP leads to more stochastic behavior, and reduced learning rate leads to less confident choices in later trials.

**Figure S2.**
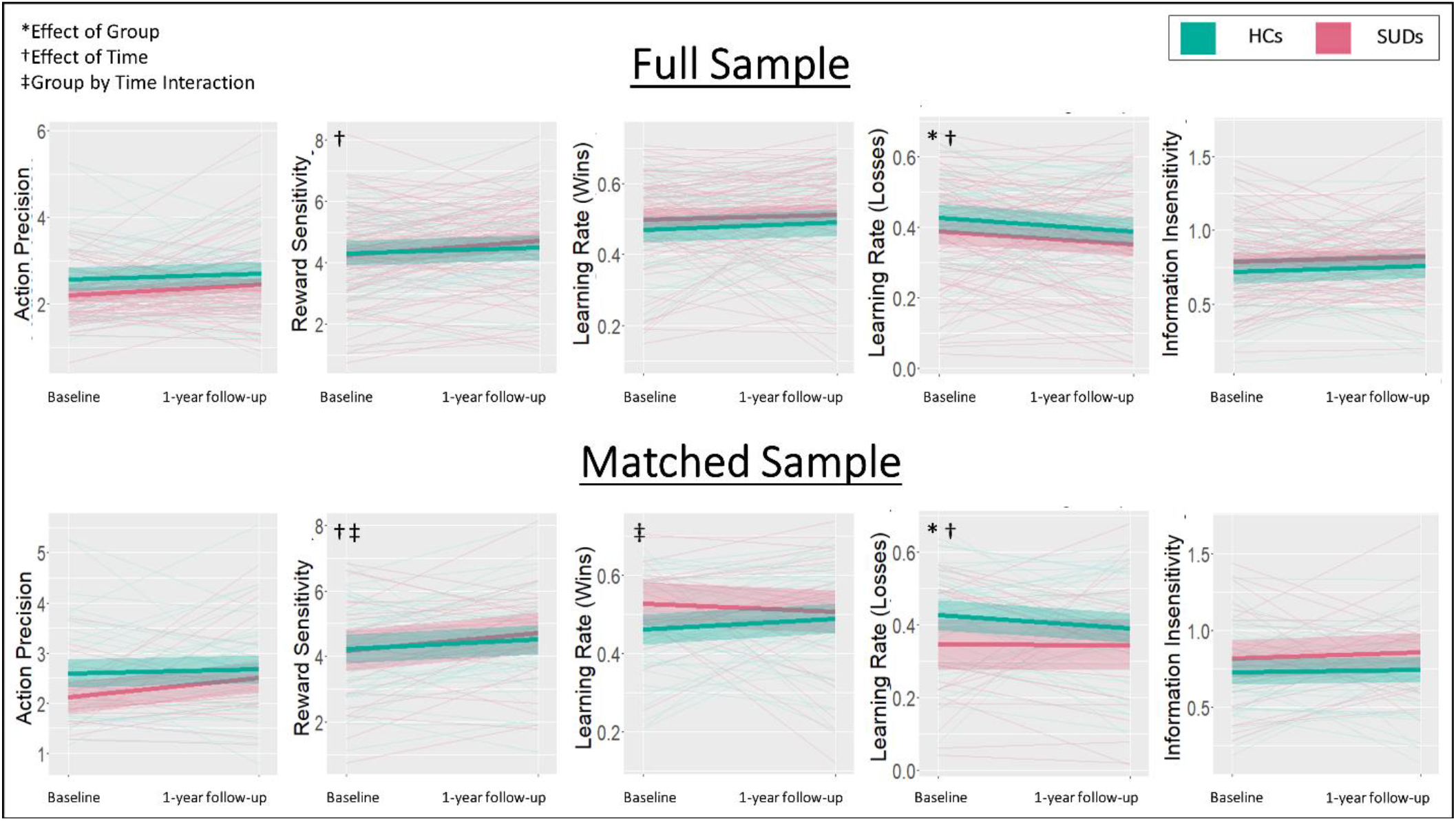
Spaghetti plots showing individual changes from baseline to follow-up, as well as group means and standard errors, for all model parameters in the full and matched samples.

**Figure S3.**
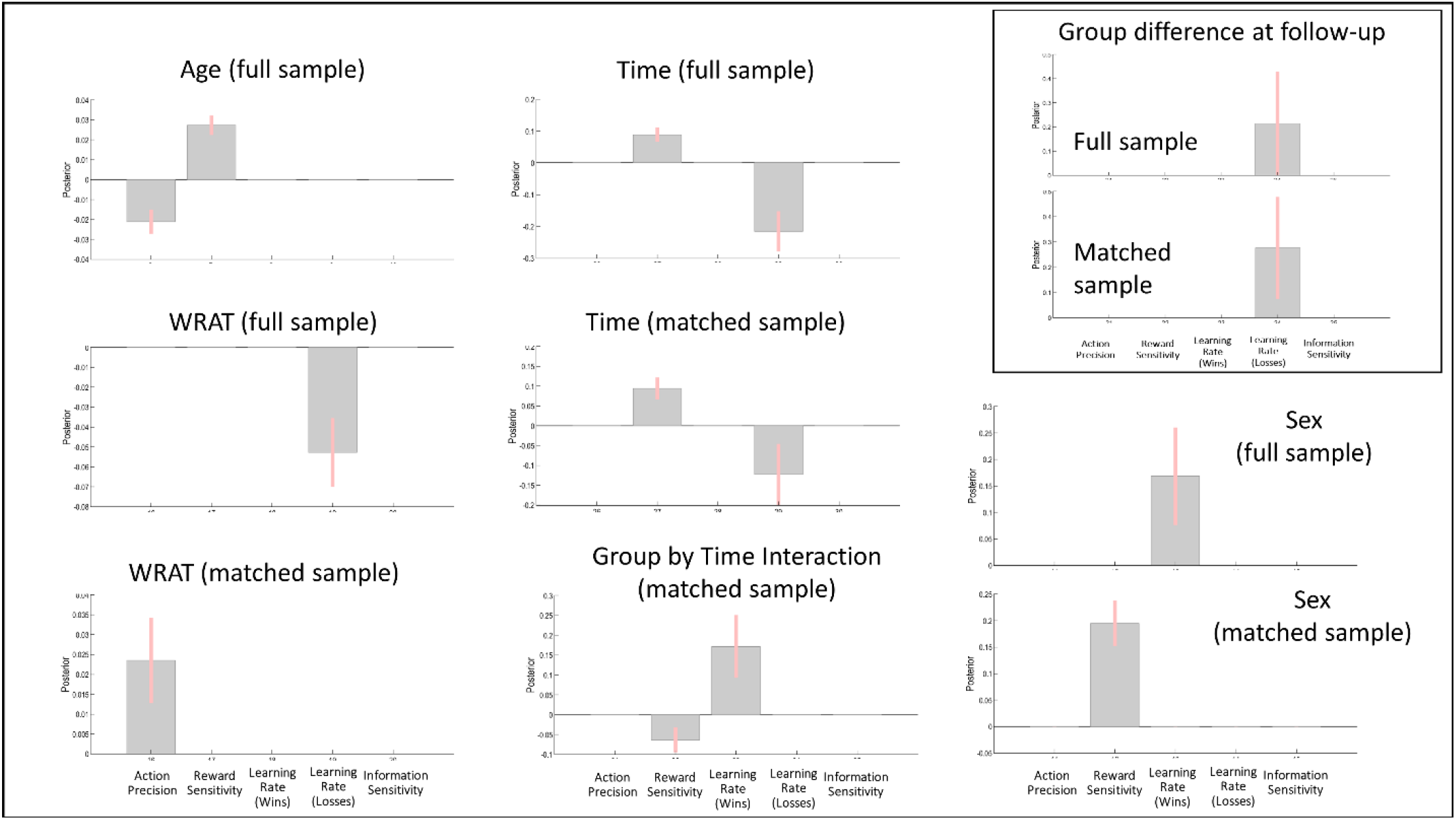
Illustration of effect sizes for additional effects found in PEB analyses that were not illustrated in the main text. For the group by time interaction, a positive value indicates increases over time in HCs and decreases over time in SUDs.

**Figure S4.**
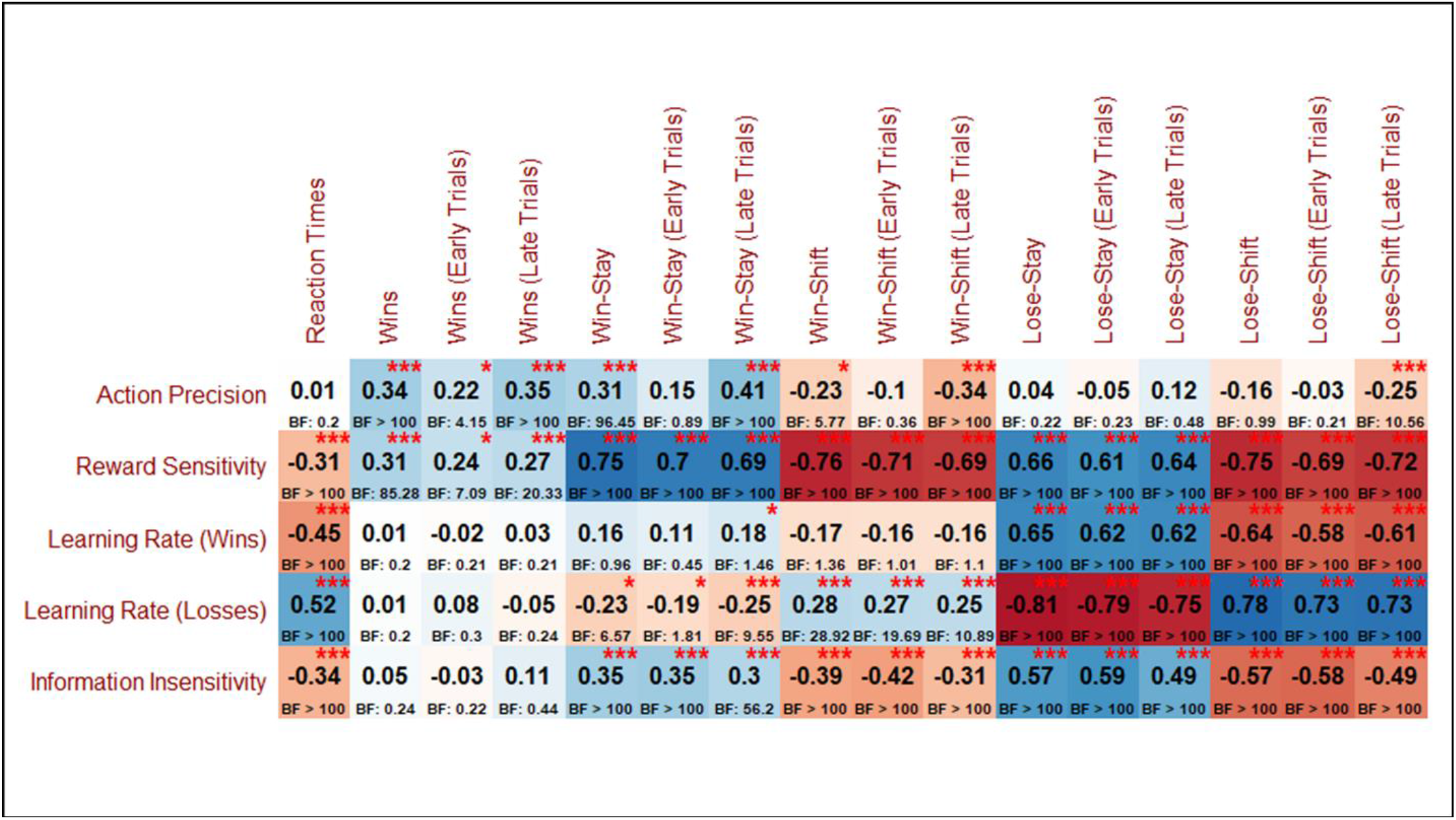
Correlations between model parameters and model-free behavior. Early trials = trials 2-7 per game. Late trials = trials 8-15 per game.

## Notes

### Competing Interest Statement

The authors have declared no competing interest.

### Clinical Trial

NCT02450240

### Author Declarations

The study was approved by the Western Institutional Review Board.

